# Relationship between blood-cerebrospinal fluid barrier integrity, cardiometabolic and inflammatory factors in schizophrenia-spectrum disorders

**DOI:** 10.1101/2024.09.17.24313817

**Authors:** Vladislav Yakimov, Iris Jäger, Lukas Roell, Emanuel Boudriot, Verena Meisinger, Mattia Campana, Lenka Krčmář, Sean Halstead, Nicola Warren, Dan Siskind, Isabel Maurus, Alkomiet Hasan, Peter Falkai, Andrea Schmitt, Florian J. Raabe, Daniel Keeser, CDP-Working Group, Elias Wagner, Joanna Moussiopoulou

**Affiliations:** Department of Psychiatry and Psychotherapy, LMU University Hospital, LMU Munich, Munich, Germany; International Max Planck Research School for Translational Psychiatry (IMPRS-TP), 80804 Munich, Germany; NeuroImaging Core Unit Munich (NICUM), LMU University Hospital, LMU Munich, 80336 Munich, Germany; Max Planck Institute of Psychiatry, 80804 Munich, Germany; Department of Psychiatry and Psychotherapy, Medical Faculty, LVR Hospital of the Heinrich Heine University Düsseldorf, Düsseldorf, Germany; Medical School, The University of Queensland, Brisbane, QLD, Australia; Metro South Addiction and Mental Health, Brisbane, QLD, Australia; Department of Psychiatry, Psychotherapy and Psychosomatics, Faculty of Medicine, University of Augsburg, 86156 Augsburg, Germany; German Center for Mental Health (DZPG), partner site Munich/Augsburg, Germany; Laboratory of Neuroscience (LIM27), Institute of Psychiatry, University of Sao Paulo, São Paulo, Brazil; Evidence-based Psychiatry and Psychotherapy, Faculty of Medicine, University of Augsburg, Stenglinstrasse 2, 86156 Augsburg, Germany

**Keywords:** schizophrenia, CNS barriers, cardiovascular system, inflammation, ventricular system

## Abstract

The blood-cerebrospinal fluid barrier (BCB) builds an integral interface between the central nervous system and the periphery and appears to be impaired in a substantial proportion of individuals with schizophrenia-spectrum disorders (SSD). Even though a disruption of the BCB is associated with higher symptom severity, factors linked to BCB disruption in SSDs have been minimally investigated.

To address this gap, 57 inpatients with SSD underwent cerebrospinal fluid (CSF) and blood analyses as well as comprehensive clinical assessments. In a subgroup of 28 participants structural magnetic resonance imaging (MRI) was performed. We developed a BCB dysfunction score, employing principal component analysis of CSF/serum albumin, CSF/serum IgG ratios and total protein levels in CSF, with higher values indicating stronger abnormalities. We calculated multiple regression models to explore the associations between BCB integrity and cardiometabolic, inflammatory, brain morphometric, and clinical measures respectively.

BCB dysfunction score was negatively associated with high-density lipoprotein cholesterol and positively associated with total cholesterol, low-density lipoprotein cholesterol, and triglycerides. Furthermore, we observed a trend towards a positive association between BCB dysfunction score and treatment resistance that did not survive multiple testing correction. We did not find significant associations between the BCB composite score and any other assessed cardiometabolic, inflammatory or cerebroventricular measures.

These findings suggest that BCB integrity is associated with dyslipidemia in SSD, highlighting the interplay between cardiometabolic risk factors and brain health in SSD. Addressing cardiometabolic health in individuals with SSD might thus have implications beyond physical health, potentially influencing the integrity of the BCB and, consequently, clinical trajectories.

## Introduction

Schizophrenia-spectrum disorders (SSD) are among the leading causes of morbidity worldwide due to high rates of treatment resistance as well as cognitive and functional impairment^1^. Compared to the general population, life expectancy is reduced by ∼15 years^2^ not only due to high rates of suicide, but also due to high prevalence of somatic comorbidities such as cardiovascular diseases^3,4^. Of note, data from drug-naïve first episode schizophrenia (SCZ) patients^4,5^, as well as genetic studies^6,7^, indicate that metabolic disturbances are not just sequelae of unhealthy lifestyle factors and adverse drug reactions, but potentially contribute to SSD pathophysiology.

Metabolic syndrome (MetS) is defined as the occurrence of at least three of interrelated cardiometabolic risk factors including central obesity, hypertension, hyperglycemia, and dyslipidemia^8^. It is highly prevalent in SSD^9^ and high-quality meta-analytic evidence suggests a link between MetS and cognitive impairment in people with SCZ^10,11^. Furthermore, a study from the ENIGMA Working Group demonstrated that body-mass-index (BMI) (as a proxy for obesity) was additively associated with structural alterations in many of the same brain regions affected in schizophrenia, including changes in cortical thickness^12^. Despite the limitations of the cross-sectional nature of this data, they suggest a complex relationship between metabolic disturbances, brain structure, and cognitive functions in individuals with SSD.

Even though the exact pathophysiology of SSD remains elusive, recent research has highlighted the role of immune system dysfunction as a contributing factor in a subset of individuals with SSD^13,14^. Converging evidence from genetic^15^, as well as, large-scale epidemiological studies^16,17^ point to the role of immune dysregulation in SSD^18^. Furthermore, preliminary evidence from multimodal studies combining neuroimaging data with peripheral inflammatory markers suggests a link between peripheral low-grade inflammation and structural, as well as, functional cerebral changes, which potentially increase the risk for more pronounced psychopathology^14^.

Impairments of the blood-brain barrier (BBB) and the blood-cerebrospinal fluid (CSF) barrier (BCB) are common findings in SSD^19,20^. While both barriers share similar functions, the BBB is spread throughout the brain whereas the BCB is mainly formed by epithelial cells of the choroid plexus (ChP) and the arachnoid membrane facing the CSF^21,22^. Of note, CSF/serum ratios of proteins such as albumin and immunoglobulin G (IgG) are technically an indirect measure of BCB and not BBB integrity^21,22^. Some of the functions of both interfaces include ensuring a stable milieu^18^, which is crucial for intact neural signaling in the brain, as well as transport of nutrients, oxygen, and waste products^18^. BCB and BBB also act as central immunological nodes, building the interface between central and peripheral immune system and coordinating access of leukocytes to the central nervous system (CNS)^18,23^. Hence, a barrier disturbance is likely to be associated with disruption of brain homeostasis and functioning with potential relevance for SSD (psycho)pathology^18^. In line with those findings, there is growing evidence suggesting increased ChP volumes^24^, variability^25^, altered ChP epithelia^26^, and associated upregulation of immune genes in the ChP^27^ of individuals with SSD. Despite the anatomical and functional relevance of the ChP for BCB^23^ and the growing evidence for alterations of both ChP^24^ and BCB^19^ in SSD, the relationship between those two has never been studied before.

Even though the exact cause of BCB disruption in SSD is not known, one of the hypotheses states that it occurs following a primary inflammatory insult^18^. Once disrupted, this might render the brain susceptible to peripheral immune effectors with the potential to disturb brain function^18^. Since blood vessels and especially endothelial cells are an integral building unit of the BCB, it is likely that vascular dysfunction, as a result of high cardiometabolic burden (e.g., hypertension, diabetes, obesity and dyslipidemia), might also be of relevance.

To address these questions, in our study we 1) explored the associations between BCB integrity and different disease characteristics (duration of illness, duration of antipsychotic treatment, first episode psychosis status) and clinical factors (global functioning, treatment resistance, positive, negative, and general symptoms, cognitive impairment). Next, we 2) studied the links between BCB integrity and peripheral inflammatory markers as well as 3) cardiometabolic risk factors. Lastly, in a subgroup of participants with SSD we 4) investigated the associations between BCB integrity and volumes of cerebroventricular regions, such as choroid plexus and lateral ventricles.

## 2. Methods

### 2.1. Participants

This study was conducted in the context of the IMPACT study, an ongoing add-on study to the Munich Mental Health Biobank (project number 18-716)^28^, and approved by the ethics committee of the Faculty of Medicine, LMU University Hospital Munich (project number 21- 1139).

The recruitment of study participants was conducted at the Department of Psychiatry and Psychotherapy, LMU University Hospital, LMU Munich, Germany, between July 26, 2018, and April 24, 2023. Only inpatients (N=57) were included. All study participants provided written informed consent and were between 18 and 65 years old. Included patients had a primary diagnosis of schizophrenia, schizoaffective disorder, delusional disorder, or brief psychotic disorder, collectively referred to as SSD throughout the manuscript.

Exclusion criteria were as follows: concurrent clinically relevant neurological disorders, such as multiple sclerosis and epilepsy, history of encephalitis, meningitis, stroke, traumatic brain injury or cerebral surgery, current pregnancy or lactation, rheumatic disorders, inflammatory bowel disease, active malignancy, and acute or chronic infection.

### 2.2. Clinical assessments

The clinical characterization was performed by trained study personnel as previously described by our working group^29,30^. The German version 7.0.2 of Mini International Neuropsychiatric Interview (M.I.N.I.)^31^, based on DSM-5 criteria, was conducted with all study participants to confirm the diagnosis. Symptom severity was assessed with the Positive and Negative Syndrome Scale (PANSS)^32^ and global functioning with the Global Assessment of Functioning (GAF)^33^ scale. The assessments were performed within four weeks around the lumbar puncture. Information regarding medication, duration of illness (DUI), BMI, blood pressure, heart rate, concomitant somatic conditions, and current smoking status was collected based on self-report and by examining medical reports. Current treatment or history of clozapine use was used as a proxy for treatment resistance, as previously suggested^34^.

To assess the cognitive performance of the participants, the Montreal Cognitive Assessment (MoCA)^35^ and the Trail-Making-Test (TMT, part A and B) were performed in a subgroup of participants.

### 2.3. Blood and cerebrospinal fluid analyses

In line with recommendations from the German schizophrenia guideline^19,36^, lumbar puncture was offered to all patients with first- (FEP) or multi-episode psychosis (MEP), who had not yet received CSF analysis in the past as part of the diagnostic work-up to exclude concurrent somatic etiologies. Paired CSF and serum samples were analyzed as part of the clinical routine diagnostics by the Institute of Laboratory Medicine, LMU Munich.

Most of the study participants underwent a basic blood test, including full blood (N = 54) and serum (N = 52 – 56, depending on the variable assessed) analyses, within 3 weeks from the lumbar puncture as part of the clinical routine in our clinic. This was done during the morning hours under fasting conditions. The full blood analysis included a complete blood count, and the serum analysis included assessment of C-reactive protein (CRP), triglycerides, total cholesterol, low-density lipoprotein (LDL) cholesterol, high-density lipoprotein (HDL) cholesterol, glycated hemoglobin (HbA1c), albumin, immunoglobulin G (IgG) levels, and the presence of oligoclonal bands (OCBs). To compute CSF/serum albumin and IgG ratios, serum and CSF were collected and assessed at the same timepoint. The neutrophil-to-lymphocyte ratio (NLR) and monocyte-to-lymphocyte ratio (MLR) were calculated by dividing the absolute number of neutrophils and monocytes each by the absolute number of lymphocytes per individual^37^. Further information is provided in the supplementary methods section.

### 2.4. Magnetic resonance imaging

A subset of 28 participants underwent brain magnetic resonance imaging (MRI) using a Siemens Magnetom Prisma 3T scanner (Siemens Healthineers AG, Erlangen, Germany) equipped with a 32-channel head coil. Regional brain volumes, including the lateral ventricles, third and fourth ventricles were quantified in cubic millimeters using FreeSurfer software (version 7.3.2; https://surfer.nmr.mgh.harvard.edu)^38^. We utilized the FreeSurfer atlas to obtain the volumes of the ventricles. Additionally, the choroid plexus (ChP) in the lateral ventricles was manually segmented on the 3D-T1 images by one of the first authors (IJ), who was trained by a neuroimaging expert (DK). We employed ITK-SNAP software, version 4.2.0 (http://www.itksnap.org). The rater was blinded regarding clinical and imaging data and followed a previously published protocol for ChP segmentation^39^. The ChP as well as the ventricle measures were adjusted for total intracranial volume using the proportions method^40^. Structural MRI data quality control was performed as previously described by our working group^41^. Further information is provided in the supplementary methods section.

### 2.5. Assessment of BCB dysfunction

To quantify the integrity of the BCB, we computed a principal component analysis (PCA) including the CSF/serum albumin ratio, CSF/serum IgG ratio, and total protein levels in CSF as variables of interest (Fig. S1). All three measures have been associated with BCB disruption and regarded as biomarkers for BCB functioning^42,43^. The variables were scaled, using the *“scale”* function in R, to increase comparability, given their differing scales. PCA reduces the dimensionality of data that are correlated^44^. Instead of analysing all three measures individually PCA summarizes the information represented by those measures in one BCB composite score, which mirrors the integrity of the BCB (the higher the BCB composite score, the lower the BCB integrity and the higher the degree of BCB dysfunction) better than any of those measures alone.

### 2.6. Statistical analyses

The R language (v4.2.1, R Core Team, 2021) in RStudio environment (RStudio Team, 2020)^45^ was used for all statistical analyses and visualizations. The following statistical tests were used to compare differences in demographic and clinical variables between SSD and HC: Fisher’s exact test for categorical variables, Welch’s *t* test for normally distributed and Wilcoxon rank-sum test for non-normally distributed continuous variables. Shapiro–Wilk test was used to assess normality within groups^46^.

To investigate the relationships between BCB composite score (predictor variable) and measures of psychopathology, cognition, or cerebroventricular measures as outcome variables we computed multiple linear regressions, controlling for age and sex as covariates. In the linear regression models including cognition measures we additionally controlled for years of education as a covariate. To investigate the association between BCB composite score (predictor variable) and treatment resistance (outcome variable), we computed a logistic regression controlling for age, sex, smoking status, and BMI as covariates. To study the relationships between cardiometabolic risk factors (e.g., total cholesterol, HbA1c) or inflammatory measures (e.g., CRP) as predictor variables and BCB composite score as outcome variable, we conducted multiple linear regressions controlling for age, sex, BMI and smoker status, as previously suggested^47,48^.

The threshold for statistical significance was set at *p* value < 0.05. Results from the descriptive statistics are shown as mean ± standard deviation (SD). We employed the Benjamini-Hochberg method^49^ for multiple testing correction within every group of sub-analyses. False discovery rate (FDR) adjusted *p* values were reported as *q* values.

## 3. Results

### 3.1. Cohort characteristics

The study cohort consisted of 57 individuals with SSD who were inpatients at the Department of Psychiatry and Psychotherapy, LMU university hospital, Munich, and underwent a lumbar puncture for diagnostic reasons. The cohort included 42 (74%) male and 15 (26%) female participants with an average age of 34.32 ± 11.97 years. Nearly half of the participants included (27/55; 49%) were active smokers. Thirty-eight individuals were diagnosed with schizophrenia (67%) and 13 with brief psychotic disorder (22%), five with schizoaffective disorder (9%), and one with delusional disorder (2%). Thirty-five (61%) of the participants had a first episode of psychosis at the time of inclusion. The mean duration of illness was 60.24 months (SD = 94.20) and the mean duration of antipsychotic treatment at the time of inclusion was 47.88 months (SD = 95.89). Forty-seven participants were assessed with the PANSS and averaged a total score of 62.87 ± 13.09. The average GAF score of the participants was 46.87 ± 11.42. Forty- one individuals performed the cognitive tests scoring an average of 25.93 ± 3.65 in the MoCA. They required an average of 33.39 ± 14.83 seconds to complete the TMT-A and 94.38 ± 65.15 seconds for the TMT-B. One of the 41 participants did not complete the TMT-B (Table 1).

**Table 1.** Cohort characteristics.

| Demographic characteristics | SSD |  |
| --- | --- | --- |
|  | *Mean ± SD* | *N* |
| Age, years | 34.32 ± 11.97 | 57 |
| BMI | 26.04 ± 4.09 | 57 |
| Education (years) | 14.86 ± 4.10 | 41 |
|  | *n (%)* |  |
| Sex, male:female | 42:15 (74%) | 57 |
| Current smoking, yes:no | 27:28 (49%) | 55 |
| Clinical characteristics | *Mean ± SD* | *N* |
| Duration of illness, months | 60.24 ± 94.20 | 56 |
| Duration of antipsychotic treatment, months | 47.88 ± 95.89 | 56 |
| PANSS positive symptoms | 15.34 ± 4.14 | 47 |
| PANSS negative symptoms | 15.60 ± 5.75 | 47 |
| PANSS general symptoms | 31.83 ± 6.56 | 47 |
| PANSS total score | 62.87 ± 13.09 | 47 |
| GAF | 46.87 ± 11.42 | 47 |
| TMT A time (seconds) | 33.39 ± 14.83 | 41 |
| TMT B time (seconds) | 94.38 ± 65.15 | 40 |
| MoCA score | 25.93 ± 3.65 | 41 |
| Systolic Blood Pressure (mmHg) | 121.50 ± 10.55 | 57 |
|  | *n (%)* |  |
| First episode psychosis, yes:no | 35:22 (61.4%) | 57 |
| Clozapine lifetime, yes:no | 7:48 (12.7%) | 55 |
| Diagnosis (*DSM-5*) | *n* (%) |  |
| Schizophrenia | 38 (66.7%) |  |
| Brief psychotic disorder | 13 (22.8%) |  |
| Schizoaffective disorder | 5 (8.8%) |  |
| Delusional disorder | 1 (1.7%) |  |

GAF, global assessment of functioning; MoCA, Montreal Cognitive Assessment; *N*, number of participants; PANSS, Positive and Negative Syndrome Scale; *SD*, standard deviation; SSD, Schizophrenia Spectrum Disorder; TMT, Trail making Test.

### 3.2. Cerebrospinal fluid and blood characteristics

Mean number of white blood cells was 0.88 ± 1.09/µl and no participant demonstrated a CSF pleocytosis. Oligoclonal IgG bands were present in 5/57 (8.8%) participants and none of these cases were of intrathecal origin (Table St1). Mean CSF protein level was 42.42 ± 18.55 mg/dl, mean CSF/serum albumin ratio (Q_alb_) was 6.53 ± 3.36 and mean CSF/serum IgG ratio was 3.18 ± 1.66. The age-adjusted Q_alb_ was elevated in 24/57 (42.1%) participants and neuronal autoantibodies were not detected in CSF or blood in any of the participants.

Following our PCA, the first principal component (PC1) explained 98.5% of the data variance (Fig. S1), so it was used to create a composite score that mirrors the BCB integrity (referred to as BCB composite score throughout the manuscript). Higher values of BCB composite score indicate higher level of disruption and lower BCB integrity.

In our cohort the mean numbers of neutrophiles, monocytes, and lymphocytes (N = 56) were 4.27 ± 1.82 thou./µl, 0.53 ± 0.17 thou./µl, and 1.86 ± 0.57 thou./µl, respectively. Mean neutrophil-to-lymphocyte ratio (NLR) was 2.43 ± 1.03, mean monocyte-to-lymphocyte ratio (MLR) was 0.30 ± 0.10 and mean CRP: 0.18 ± 0.26 mg/dl. An elevated CRP level (> 0.5 mg/dl) was found in 5/56 (8.9 %) participants. Mean total cholesterol was 178.20 ± 32.64 mg/dl, mean high-density lipoprotein (HDL) cholesterol was 53.24 ± 16.48 mg/dl, mean low-density lipoprotein (LDL) cholesterol was 109.10 ± 37.74 mg/dl. Those parameters were abnormal in 14/55 (25.5%), 27/55 (49.1%), and 19/55 (34.5%) participants respectively (Table St2). Mean triglyceride level was 118.50 ± 82.99 mg/dl, mean glycated hemoglobin (HbA1c) was 5.34 ± 0.51 %. They were elevated in 14/55 (25.5%) and 3/54 (5.6%) participants respectively.

### 3.3. Association between blood-cerebrospinal fluid barrier integrity and clinical phenotype

First, we investigated whether there was an association between psychopathology or level of functioning and BCB integrity. We found no significant associations of BCB composite score with PANSS positive, negative, general, and total scores or GAF scores (Fig. 1B, 1C, Fig. S2, Table S2). However, there was a nominally significant association between current or past treatment with clozapine and higher BCB composite score (Fig. 1A), which did not survive correction for multiple testing (estimate [95% CI] = 0.668 [0.158, 1.177]; p = 0.031; *q* = 0.186). After correcting for age, sex, and years of education, we found no significant associations between TMT A, TMT B, MoCA scores and BCB composite score (Fig 1D, Fig. S2, Table S3). Furthermore, there was no significant association between BCB composite score and duration of illness, duration of antipsychotic treatment, or first episode psychosis status (Fig. S3, Table S1).

**Figure 1.**
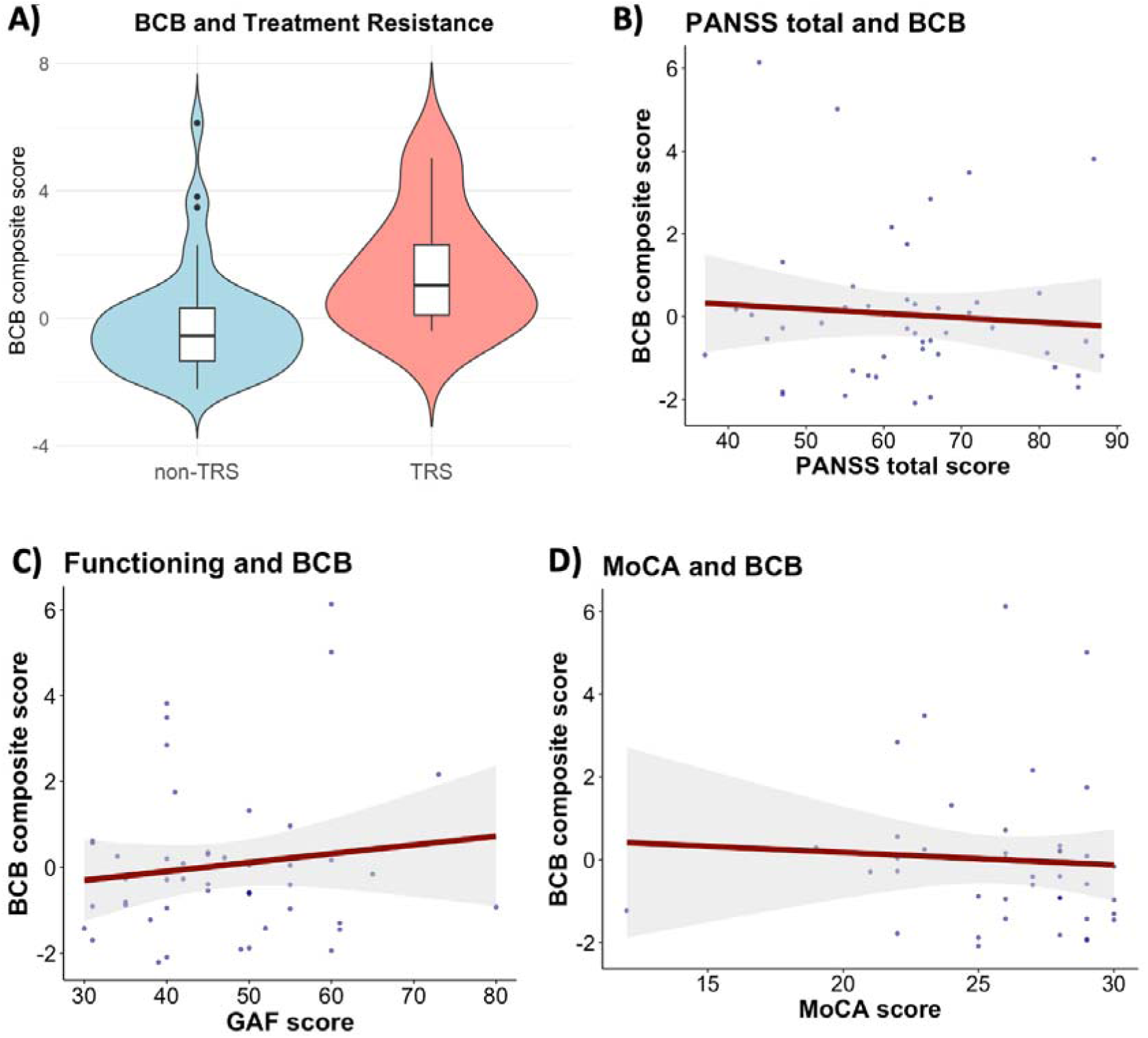
Relationship between blood-cerebrospinal fluid barrier integrity and clinical phenotype. **(A)** Comparison of mean blood-cerebrospinal fluid barrier composite score between treatment-resistant schizophrenia group (red) and non-treatment-resistant schizophrenia group (turquoise) illustrated with box and violin plots. Groups were compared using a logistic regression model, controlling for age, sex, and BMI. N_non-TRS_ = 48, N_TRS_ = 7. Regression plots illustrating associations between blood-cerebrospinal fluid barrier composite score, **(A)** PANSS total score (N = 47), **(B)** GAF score (N = 47) and **(C)** MoCA score (N = 35). Multiple linear regression models were employed, controlling for covariates. Abbreviations: N, number of participants; BCB, blood-cerebrospinal fluid barrier; TRS, treatment-resistant schizophrenia; non-TRS, non-treatment-resistant schizophrenia; PANSS, Positive and Negative Syndrome Scale; GAF, Global Assessment of Functioning scale; MoCA, Montreal Cognitive Assessment scale.

### 3.4. Relationship between cardiometabolic factors and blood-cerebrospinal fluid barrier integrity

Subsequently, we explored the relationships between cardiometabolic risk factors and the blood-cerebrospinal fluid barrier composite score in SSD participants. We found significant positive associations between total cholesterol (estimate [95% CI] = 0.026 [0.014, 0.038]; *q* = 0.003), LDL cholesterol (estimate [95% CI] = 0.023 [0.013, 0.033]; *q* = 0.001), triglycerides (estimate [95% CI] = 0.013 [0.008, 0.017]; *q* = 0.00006) on the one hand, and BCB composite score on the other as well as a significant negative association between HDL cholesterol (estimate [95% CI] = -0.045 [-0.067, -0.022]; *q* = 0.004) and BCB composite score (Figure 2, Table S4), after controlling for the covariables age, sex, BMI, and smoking status. We did not find a significant association between other cardiovascular factors such as systolic blood pressure or HbA1c, and BCB composite score (Figure S4, Table S4).

**Figure 2.**
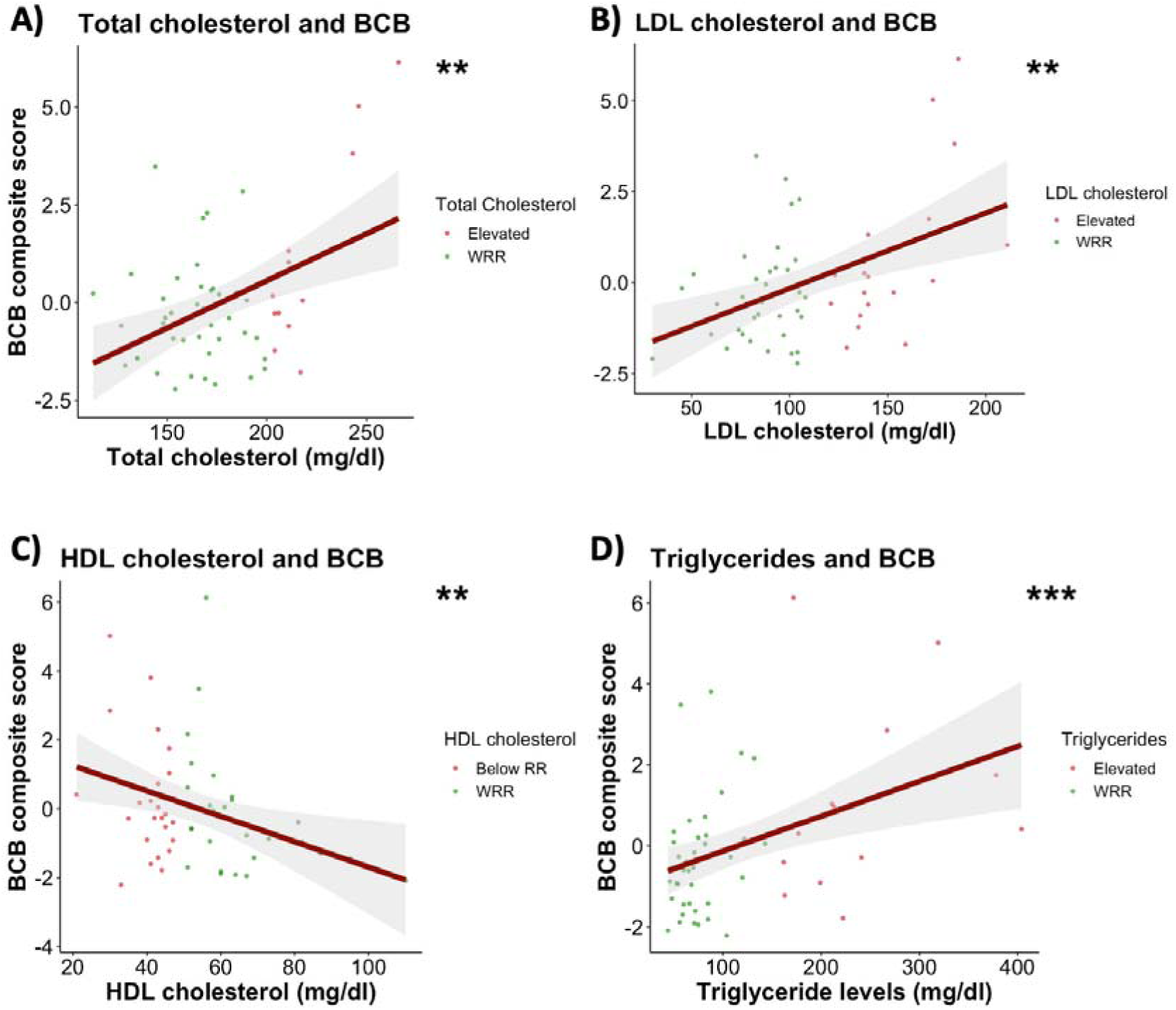
Association between blood-cerebrospinal fluid barrier integrity and serum lipids. Regression plots illustrating associations between blood-cerebrospinal fluid barrier composite score, **(A)** total cholesterol, **(B)** LDL cholesterol, **(C)** HDL cholesterol and **(D)** triglycerides. Multiple linear regression models were employed, controlling for age, sex, smoking status, and BMI. N = 53. Abbreviations: N, number of participants; BCB, blood-cerebrospinal fluid barrier; LDL cholesterol, low-density lipoprotein cholesterol; HDL cholesterol, high-density lipoprotein cholesterol; WRR, within reference range.

### 3.5. Relationship between peripheral inflammation and blood-cerebrospinal fluid barrier integrity

Next, we investigated a possible association between peripheral inflammatory factors and BCB integrity. We did not find significant relationship between absolute neutrophil, monocyte and lymphocyte counts and the BCB composite score respectively. There was also no significant association between NLR, MLR, CRP levels and BCB composite score (Fig. 3, Fig. S5, Table S5).

**Figure 3.**
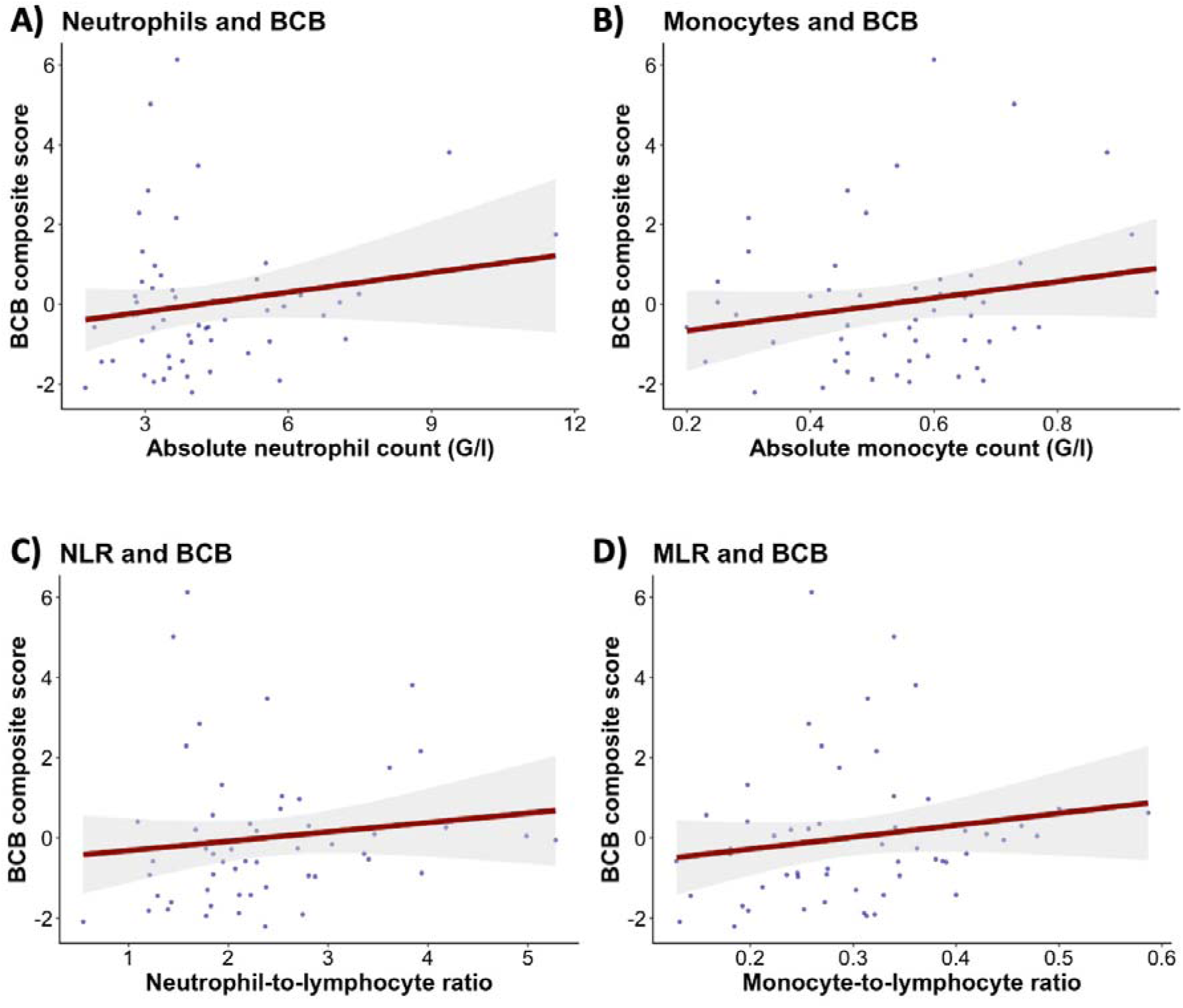
Relationship between blood-cerebrospinal fluid barrier integrity and peripheral inflammatory measures. Regression plots illustrating associations between blood-cerebrospinal fluid barrier composite score, **(A)** neutrophil count, **(B)** monocyte count, **(C)** neutrophil-to-lymphocyte ratio and **(D)** monocyte-to-lymphocyte ratio. Multiple linear regression models were employed, controlling for age, sex, smoking status, and BMI. N = 54. Abbreviations: N, number of participants; BCB, blood-cerebrospinal fluid barrier; NLR, neutrophil-to-lymphocyte ratio; MLR, monocyte-to-lymphocyte ratio.

### 3.6. Relationship between blood-cerebrospinal fluid barrier integrity and cerebroventricular regions

Due to the functional relationship between the BCB and the cerebroventricular regions, particularly the choroid plexus, we investigated the association between the level of BCB disruption and the volumes of lateral ventricles, 3^rd^ ventricle, 4^th^ ventricle and choroid plexus in a subset of participants. We found no significant associations between the BCB composite score and the volumes of the 3^rd^ ventricle, the 4^th^ ventricle, the left and right lateral ventricle or the left and right choroid plexus (Fig. 4, S6, Table S6).

**Figure 4.**
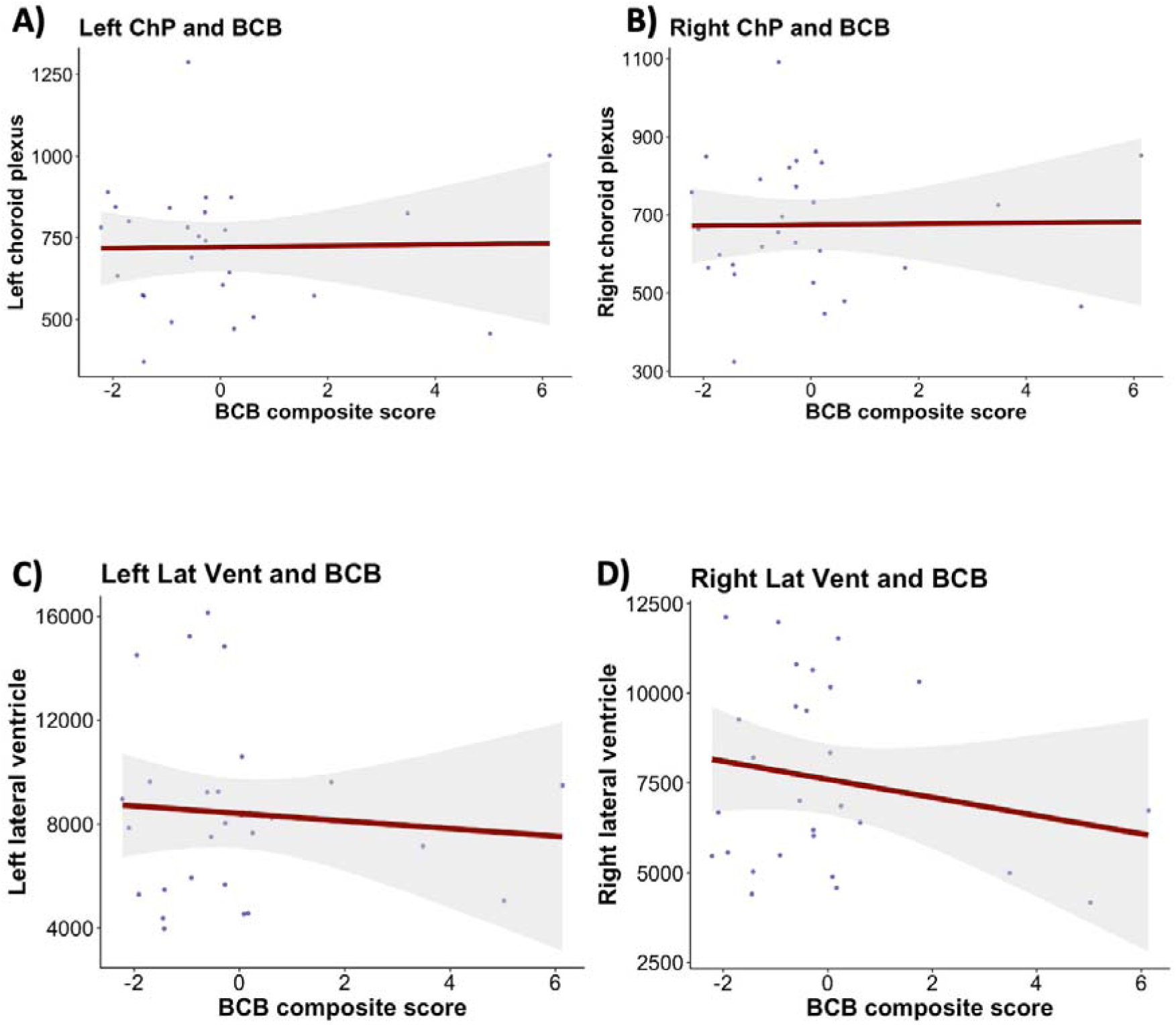
Relationship between blood-cerebrospinal fluid barrier integrity and cerebroventricular regions. Regression plots illustrating associations between blood-cerebrospinal fluid barrier composite score, **(A)** left choroid plexus volume, **(B)** right choroid plexus volume, **(C)** left lateral ventricle volume and **(D)** right lateral ventricle volume. Multiple linear regression models were employed, controlling for age and sex. N = 28. Abbreviations: N, number of participants; BCB, blood-cerebrospinal fluid barrier; Left ChP, left choroid plexus; Right ChP, right choroid plexus; Left Lat Vent, left lateral ventricle; Right Lat Vent, right lateral ventricle.

## 4. Discussion

In our study we found significant associations between higher total cholesterol, LDL cholesterol, triglycerides and disrupted BCB integrity as well as a significant association between lower HDL cholesterol and disrupted BCB integrity in individuals with SSD. We did not find significant associations between BCB composite score and measures of psychopathology or disease characteristics, despite a trend towards higher degree of BCB disruption in individuals with history of clozapine treatment. Additionally, there was no significant relationship between BCB disruption and peripheral immune markers or volumetric measures of cerebroventricular regions, respectively.

BCB disruption is a common finding in SSD^19^, but data on its relevance regarding psychopathology and disease course remains scarce. Even though some previous studies have investigated the link between abnormal CSF/serum albumin ratio (a proxy for BCB impairment) and different measures of symptomatology such as cognitive deficits^50^, positive, negative or general symptoms, no significant associations could be found across both sexes.

Interestingly, Oviedo-Salcedo, Wagner et al. found a trend towards associations between elevated CSF protein, CSF/serum albumin ratio and history of treatment with clozapine (a proxy for TRS), that did not reach statistical significance^51^. Despite the weak evidence, it aligns with our results indicating higher degree of BCB disruption in individuals with history of clozapine treatment. This data must be regarded as preliminary and interpreted cautiously (not only due to lack of significance after multiple comparison correction, but also due to the low number of individuals with history of clozapine treatment (N = 7). A recent individual participant data meta-analysis from our working group^43^ demonstrated that male individuals with SSD and elevated CSF/serum albumin ratio have significantly higher PANSS positive scores than male individuals with SSD and CSF/serum albumin ratio within the reference range. Of note, clozapine is usually prescribed to patients with treatment-resistant positive symptoms, which aligns well with these data. Overall, the results from our study and others suggest that BCB impairment might be associated with higher degree of symptom severity and even treatment resistance but should be interpreted carefully in light of the discussed limitations.

Dyslipidemia occurs in a substantial proportion of individuals with SSD^9^. To the best of our knowledge, this is the first study to show a significant relationship between blood lipid levels and BCB integrity in SSD. In line with our findings, a previous study found that subjects with Alzheimer’s disease and elevated CSF/serum albumin ratio (a proxy for BCB impairment) had significantly higher mean plasma triglycerides and lower mean HDL cholesterol than individuals without BCB impairment^52^. Preclinical evidence also points to interactions between CNS barriers and peripheral lipids^53^. For example, LDLr-knock-out mice, fed a high cholesterol diet, were more susceptible to blood-brain barrier damage and cognitive deficits^54^. In two other studies dietary-fat-induced blood-brain barrier dysfunction was restored via treatment with statins^55,56^ or ibuprofen^56^. It has been suggested that lipids affect structure and permeability of CNS barriers via altering the brain endothelial cells^53^. Indeed, the effects of dyslipidemia on blood vessels and particularly its contribution to atherosclerosis is well-studied^57^. Although we could not investigate the directionality or possible mechanisms of the association between blood lipid levels and BCB disruption, it is possible that dyslipidemia in individuals with SSD damages the endothelial cells of the vessels, forming the BCB, thus affecting its integrity. A previous study investigating the impact of hypercholesterolemia on choroid plexus epithelial cells in rabbits could demonstrate that cholesterol insults from the circulation induce dysfunction of choroid plexus epithelial cells^58^. This is particularly interesting since the choroid plexus epithelial cells are an integral building unit of the BCB^21^.

Previous studies in rats and humans have suggested that hypertension can disrupt the BCB or blood-brain barrier (BBB) integrity^59,60^ and hyperglycemia can exacerbate BBB disruption^61^, potentially through inflammatory pathways^62^. In our study, we did not find significant associations between systolic blood pressure, glycated hemoglobin and BCB composite score respectively, despite observing a trend towards positive associations. However, even though prevalence of cardiovascular diseases is increased in individuals with SSD, most of our participants did not have overt diabetes or hypertension. Thus, follow-up studies in cohorts that include more patients with cardiovascular comorbidities are needed. Furthermore, we only included systolic blood pressure from a single measurement, which could vary substantially and be influenced by multiple factors.

Building on evidence from multiple sclerosis^63^ and Alzheimer’s research^64^ as well as on the inflammatory hypothesis for schizophrenia^1^, some authors have suggested that CNS barrier disruption arises as a consequence of inflammatory insult and/or subtle immune dysregulation^65^. Even though abnormal inflammatory markers are evident in individuals with SSD both in plasma^13^ and CSF^42^, a previous study trying to link peripheral inflammation (CRP) to BCB impairment failed^19^. In our current study we also did not find significant associations between peripheral leukocytes, CRP and BCB integrity, confirming previous evidence. It is possible that markers of inflammation (non-high-sensitive CRP and immune cell counts) used in both studies were not sensitive enough to detect subtle immune dysregulation and its link to BCB impairment.

Anatomically, the BCB is formed by epithelial cells of the choroid plexus, fenestrated blood vessels and the arachnoid membrane facing the CSF^22^. Interestingly, emerging evidence points to morphological alterations of the choroid plexus in psychosis^24^ and ventricular enlargement is a well-known phenomenon in some individuals with SSD^66^. Thus, these cerebroventricular regions and BCB integrity seem to be affected in individuals with SSD; however, to the best of our knowledge, the relationship between these variables has never been investigated before in any mental disorder. In a recent study of individuals with amyotrophic lateral sclerosis the authors showed higher choroid plexus volumes compared to healthy controls and found a significant positive correlation between choroid plexus volume and CSF/serum albumin ratio (as a proxy for BCB disruption)^67^. In our study of people with SSD, we found no associations between BCB integrity and the choroid plexus or any of the ventricles.

The limitations of our study include its cross-sectional design which does not allow us to study disease progression or treatment response/remission, in relation to BCB integrity. Furthermore, a relevant part of the study participants was treated with antipsychotics, potentially influencing BCB integrity^68^. It is not clear whether treatment resistance per se or clozapine-related side effects such as dyslipidemia drive the observed association between history of clozapine treatment and BCB dysfunction. Another important limitation includes the fact that blood for immunological and cardiometabolic analyses was taken within a three-week period around the lumbar puncture. Consequently, we might have missed associations between increased BCB permeability and peripheral measures which are not stable over time, such as immune cell counts or CRP. Even though this is the first study to investigate such an association in people with SSD, the sample size was relatively small (N = 28) and subsequent well-powered studies need to address this question.

Our study adds to the growing body of literature pointing to the relevance of brain-body interactions and CNS barrier impairment for the pathophysiology of SSD. Consequently, addressing cardiometabolic factors in individuals with SSD might have implications that extend beyond physical health and influence the brain as well as the course of the disease.

Future investigations with sufficiently powered cohorts, deeper immunometabolic phenotyping performed in both blood and CSF, and longitudinal designs might help elucidate the aetiology and clinical relevance of BCB disruption in individuals with SSD.

## Data Availability

All data produced in the present study are available upon reasonable request to the authors.

## Acknowledgements

The study was endorsed by the Federal Ministry of Education and Research (Bundesministerium für Bildung und Forschung [BMBF]) within the initial phase of the German Center for Mental Health (DZPG) (grant: 01EE2303C to AH, and 01EE2303A, 01EE2303F to PF, AS). This research was supported by BMBF with the EraNet project GDNF UpReg (01EW2206) to VY and PF. The procurement of the Prisma 3T MRI scanner was supported by the Deutsche Forschungsgemeinschaft (DFG, INST 86/1739-1 FUGG). No funding was received by commercial or not-for-profit sectors. IJ and VM were supported by doctoral scholarships from the Faculty of Medicine, LMU Munich, Munich, Germany. LK, and IM were supported by the Else Krolner-Fresenius Foundation for the Residency/PhD track of the International Max Planck Research School for Translational Psychiatry (IMPRS-TP), Munich, Germany. FJR, MC, JM were supported by the Förderprogramm für Forschung und Lehre (FöFoLe) of the Faculty of Medicine, LMU Munich, Munich, Germany.

## Contributions

EW, JM and DK designed and conceptualized the IMPACT Study. VY, IJ, JM and EW designed this study and wrote the protocol. VY, JM, IJ, LK, EB, EW, and MK recruited patients and collected study data. EW trained staff on diagnostic and clinical assessments. MRI measurements were performed by JM and MK under the supervision of DK and LR. Statistical analyses were performed by VY and IJ. Data visualization was performed by VY. VY and IJ wrote the first draft of the manuscript. JM, LR, MM, EB, GH, MC, SH, NW, DS, SP, IM, AH, PF, AS, FJR, DK and EW provided critical review. VY and IJ prepared the final manuscript version with the help of all authors.

---

**Figure S1.**
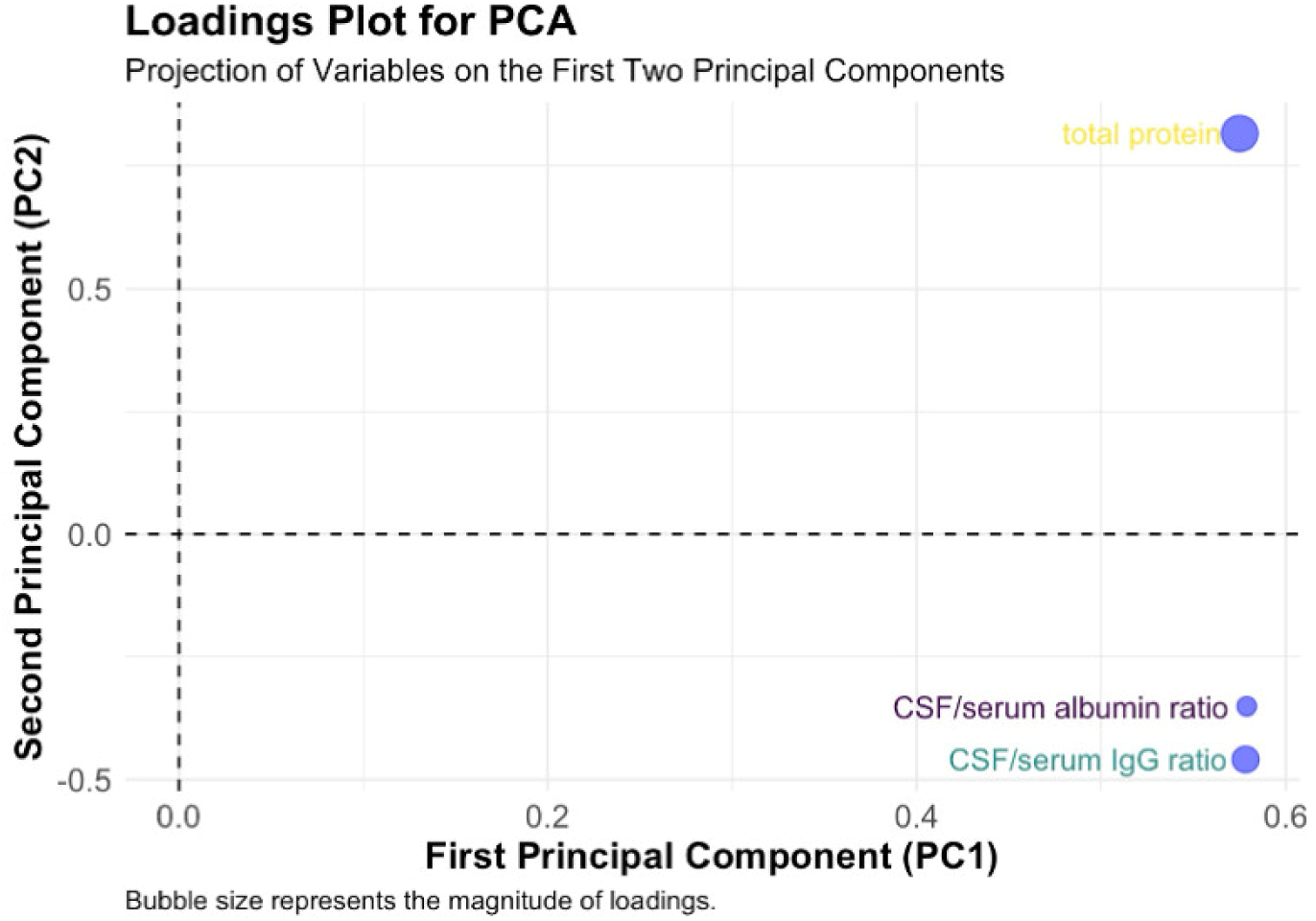

**Figure S2.**
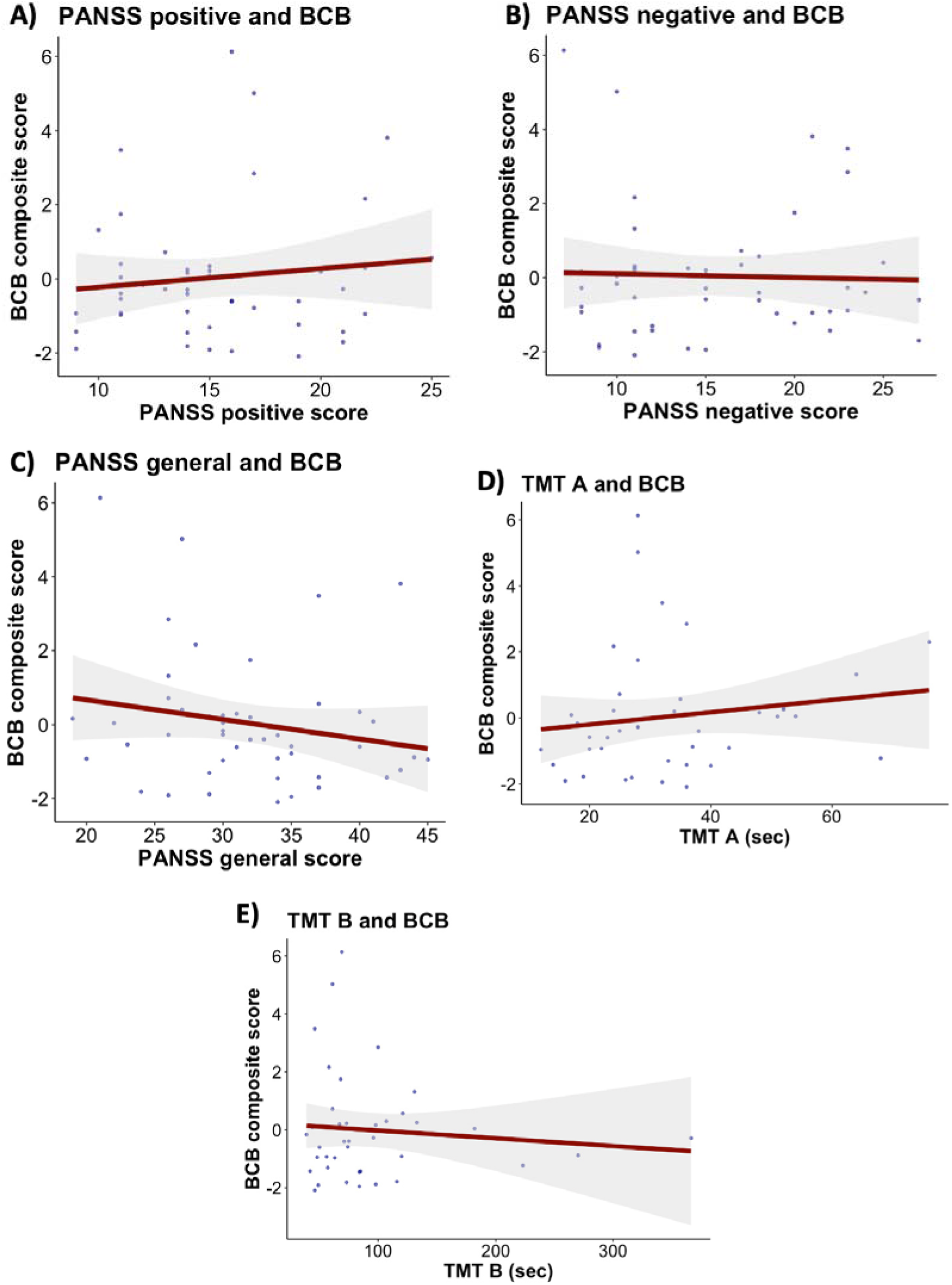

**Figure S3.**
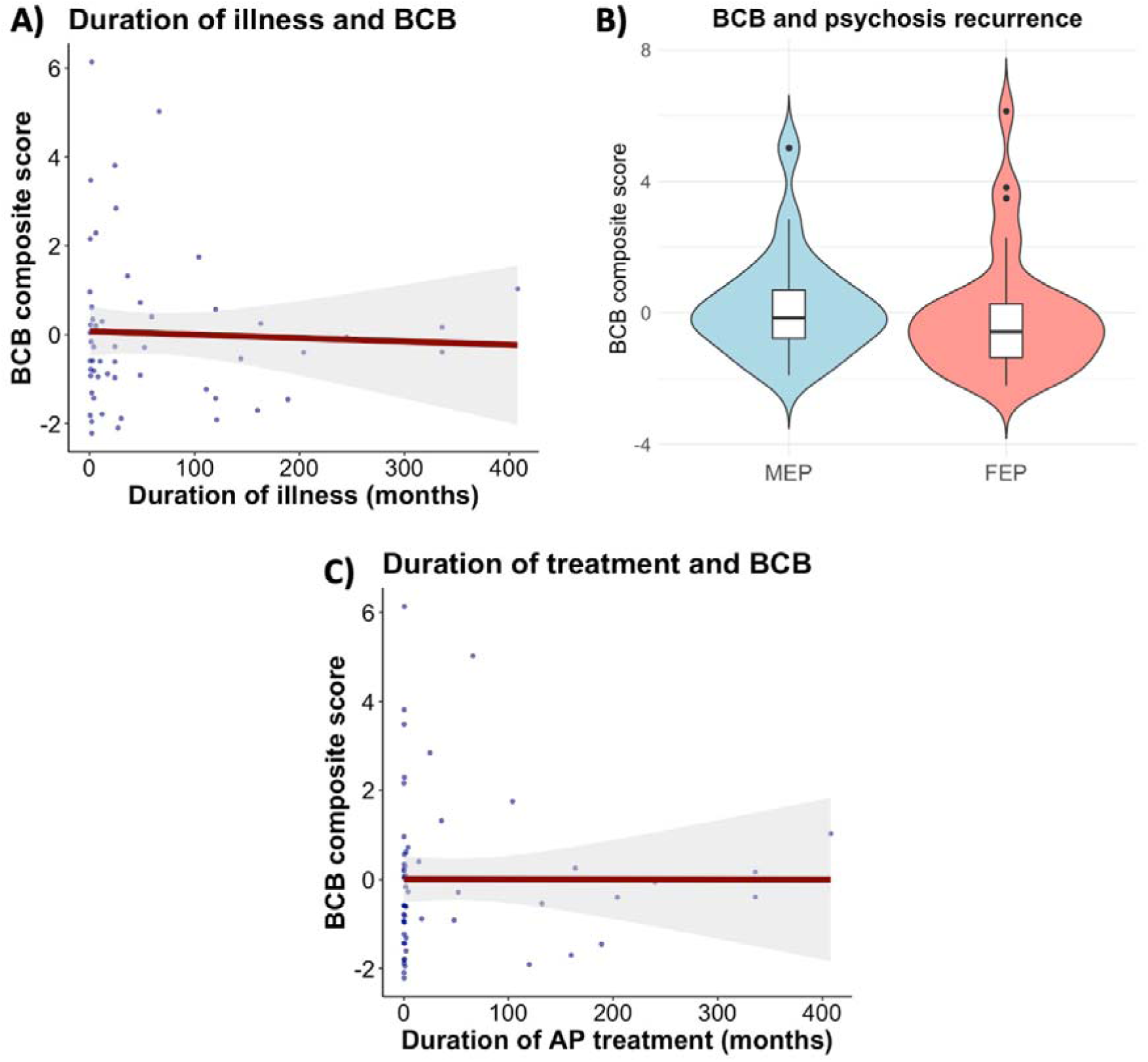

**Figure S4.**
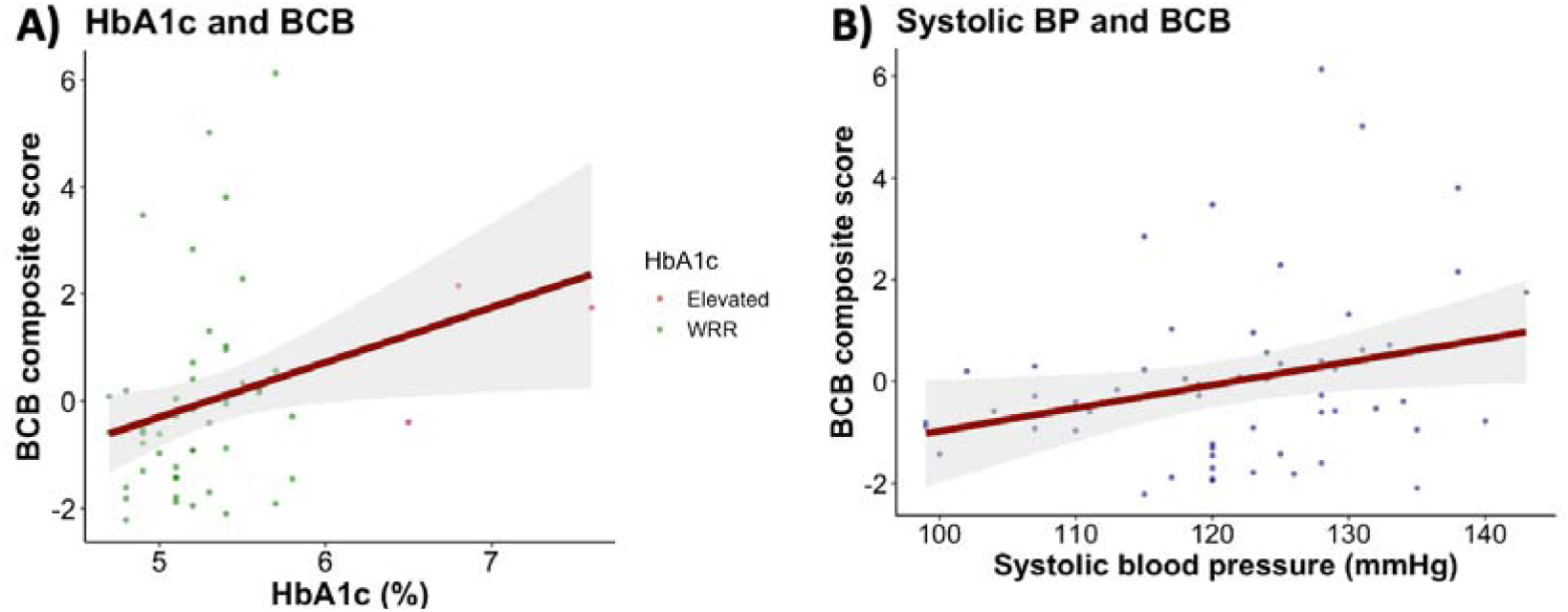

**Figure S5.**
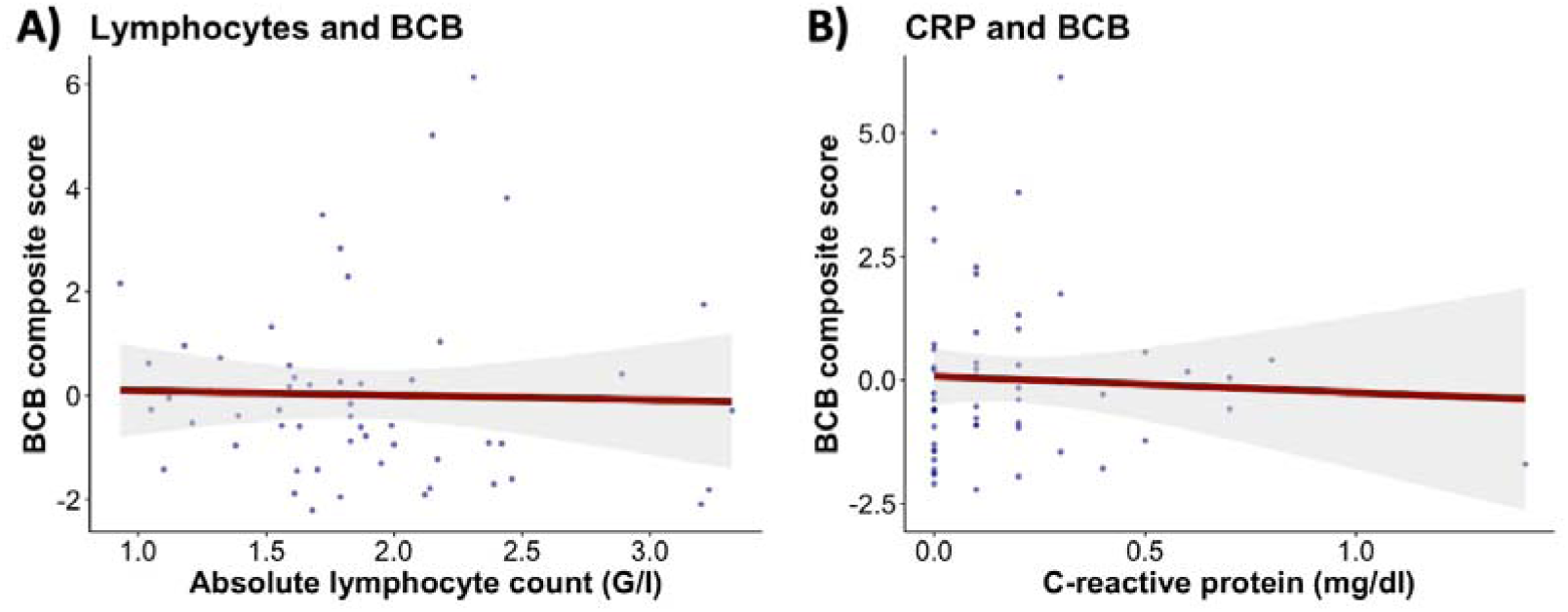

**Figure S6.**
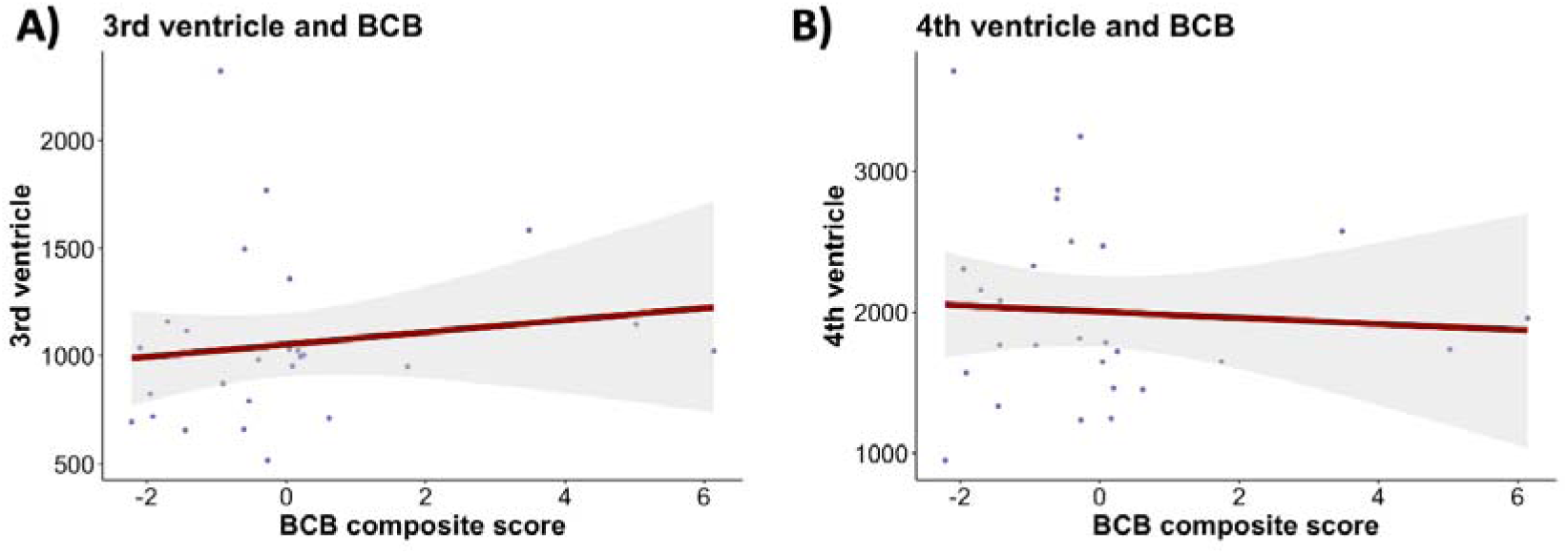

