## Supplemental File for "Relationship between blood-cerebrospinal fluid barrier integrity, cardiometabolic and inflammatory factors in schizophrenia-spectrum disorders"

Vladislav Yakimov^1,2,3#*^ (ORCID iD: 0000-0001-9559-7492), Iris Jäger^1*^, Lukas Roell^1,3^ (ORCID iD: 0000-0002-0284-2290), Emanuel Boudriot^1,4^ (ORCID iD: 0000-0001-6083-6318), Verena Meisinger^1^, Mattia Campana^1,5^, Lenka Krčmář^1,3^, Sean Halstead^6,7^ (ORCID iD: 0009-0000-4890-3506), Nicola Warren^6,7^ (ORCID iD: [0000-0002-0805-1182](https://orcid.org/0000-0002-0805-1182)), Dan Siskind^6,7^ (ORCID iD: [0000-0002-2072-9216](https://orcid.org/0000-0002-2072-9216)), Isabel Maurus^1,2^ (ORCID iD: 0000-0002-6208-5180), Alkomiet Hasan^8,9^, Peter Falkai^1,4,9^ (ORCID iD: 0000-0003-2873-8667), Andrea Schmitt^1,9,10^ (ORCID iD: 0000-0002-5426-4023), Florian J. Raabe^1,4,9^ (ORCID iD: 0000-0001-8538-0783)), Daniel Keeser^1,3^ (ORCID iD): 0000-0002-0244-1024), CDP-Working Group^1,2,3,4,5,8,9,10,11^, Elias Wagner^8,11*^, Joanna Moussiopoulou^1,3*^ (ORCID iD: 0000-0002-0157-6197)

^1^Department of Psychiatry and Psychotherapy, LMU University Hospital, LMU Munich, Munich, Germany

^2^International Max Planck Research School for Translational Psychiatry (IMPRS-TP), 80804 Munich, Germany

^3^NeuroImaging Core Unit Munich (NICUM), LMU University Hospital, LMU Munich, 80336 Munich, Germany

^4^Max Planck Institute of Psychiatry, 80804 Munich, Germany

^5^Department of Psychiatry and Psychotherapy, Medical Faculty, LVR Hospital of the Heinrich Heine University Düsseldorf, Düsseldorf, Germany

^6^Medical School, The University of Queensland, Brisbane, QLD, Australia

^7^Metro South Addiction and Mental Health, Brisbane, QLD, Australia

^8^Department of Psychiatry, Psychotherapy and Psychosomatics, Faculty of Medicine, University of Augsburg, 86156 Augsburg, Germany

^9^German Center for Mental Health (DZPG), partner site Munich/Augsburg, Germany

^10^Laboratory of Neuroscience (LIM27), Institute of Psychiatry, University of Sao Paulo, São Paulo, Brazil

^11^Evidence-based Psychiatry and Psychotherapy, Faculty of Medicine, University of Augsburg, Stenglinstrasse 2, 86156 Augsburg, Germany

* These authors contributed equally

**#Corresponding author:**

Dr. med. Vladislav Yakimov

Address: Department of Psychiatry and Psychotherapy, University Hospital, LMU Munich, Nussbaumstrasse 7, 80336 Munich, Germany

**Running title:** Blood-cerebrospinal fluid barrier disruption in schizophrenia

**Supplementary methods**

***Clinical assessments***

The clinical characterization was performed by trained study personnel as previously described by our working group^1,2^. The German version 7.0.2 of Mini International Neuropsychiatric Interview (M.I.N.I.)^3^, based on DSM-5 criteria, was conducted with all study participants to confirm the diagnosis. Symptom severity was assessed with the Positive and Negative Syndrome Scale (PANSS)^4^ and global functioning with the Global Assessment of Functioning (GAF)^5^ scale. The assessments were performed within four weeks around the lumbar puncture. Information regarding medication, duration of illness (DUI), BMI, blood pressure, heart rate, concomitant somatic conditions, and current smoking status was collected based on self-report and by examining medical reports. Current treatment or history of clozapine use was used as a proxy for treatment resistance, as previously suggested^6^.

To assess the cognitive performance of the participants, the Montreal Cognitive Assessment (MoCA)^7^ and the Trail-Making-Test (TMT, part A and B) were performed in a subgroup of participants. The MoCA assesses the following cognitive domains: short-term memory, visuospatial abilities, executive functions, attention, concentration, working memory, orientation, and language. It has been validated as a practical tool for measuring cognitive deficits in individuals with SSD^8^. The TMT covers the cognitive domains attention, visual search and scanning, processing speed, task switching, cognitive flexibility and executive function^9^, which are often affected in individuals with SSD.

***Blood and cerebrospinal fluid analyses***

In line with recommendations from the German schizophrenia guideline^10,11^, lumbar puncture was offered to all patients with first- (FEP) or multi-episode psychosis (MEP), who had not yet received CSF analysis in the past as part of the diagnostic work-up to exclude concurrent somatic etiologies. Paired CSF and serum samples were analyzed as part of the clinical routine diagnostics by the Institute of Laboratory Medicine, LMU Munich.

The CSF analysis (Table St1) included white blood cell counts (ref.: ≤ 5), total protein (ref.: 15-45 mg/dl), albumin (ref.: 0.1 – 0.3 g/l), and immunoglobulin G (IgG) (ref.: ≤ 0.034 g/l) levels as well as the presence of oligoclonal IgG bands (OCBs) and neuronal antibodies (incl., N-methyl-D-aspartate receptor (NMDA) antibodies, α-amino-3-hydroxy-5-methyl-4-isoxazolepropionic acid (AMPA) 1/2 receptor antibodies, dipeptidyl-peptidase-like protein 6 (DPPX) antibodies, contactin-associated protein-like 2 (CASPR2) antibodies, leucine-rich glioma-inactivated 1 (LGI1) antibodies, gamma-aminobutyric acid B receptor (GABA B) 1/2 antibodies).

Most of the study participants underwent a basic blood test, including full blood (N = 54) and serum (N = 52 – 56, depending on the variable assessed) analyses, within 3 weeks from the lumbar puncture as part of the clinical routine in our clinic. This was done during the morning hours under fasting conditions. The full blood analysis included a complete blood count and the serum analysis included assessment of C-reactive protein (CRP) (ref.: ≤ 0.5 mg/dl), triglycerides (ref.: ≤ 150 mg/dl), total cholesterol (ref.: ≤ 200 mg/dl), low-density lipoprotein (LDL) cholesterol (ref.: ≤ 116 mg/dl), high-density lipoprotein (HDL) cholesterol (ref.: > 40 mg/dl), glycated hemoglobin (HbA1c) (ref.: ≤ 5.7%), albumin (ref.: 3.5 – 5.2 g/dl), IgG levels (ref.: 7 – 16 g/l), and the presence of OCBs. Oligoclonal IgG bands in CSF and serum (collected at the same timepoint) were detected using a SEBIA HYDRASYS 2 SCAN FOCUSING semiautomated instrument according to the manufacturer`s instructions, which enables immunofixation and direct detection of OCBs on agarose gels using the HYDRAGEL 9CSF kit^10^. Cut-off values of CSF/serum albumin ratios (Q_alb_) were adjusted to age with the formula: Q_alb_ = (4 + age/15) × 10^–3 12^. To compute CSF/serum albumin and IgG ratios, serum and CSF were collected and assessed at the same timepoint. The neutrophil-to-lymphocyte ratio (NLR) and monocyte-to-lymphocyte ratio (MLR) were calculated by dividing the absolute number of neutrophils and monocytes each by the absolute number of lymphocytes per individual^13^.

***Magnetic resonance imaging***

A subset of 28 participants underwent brain magnetic resonance imaging (MRI) using a Siemens Magnetom Prisma 3T scanner (Siemens Healthineers AG, Erlangen, Germany) equipped with a 32-channel head coil. T1-weighted scans were acquired using a magnetization-prepared rapid gradient echo (MP-RAGE) sequence with an isotropic voxel size of 0.8 mm^3^, 208 slices, repetition time of 2500 ms, echo time of 2.22 ms, flip angle of 8°, and a field of view of 256 mm. Regional brain volumes, including the lateral ventricles, third and fourth ventricles were quantified in cubic millimeters using FreeSurfer software (version 7.3.2; <https://surfer.nmr.mgh.harvard.edu>)^14^. We utilized the FreeSurfer atlas to obtain the volumes of the left and right lateral ventricles, the third ventricle and the fourth ventricle. Additionally, the choroid plexus (ChP) in the lateral ventricles was manually segmented on the 3D-T1 images by one of the first authors (IJ), who was trained by a neuroimaging expert (DK). We employed ITK-SNAP software, version 4.2.0 (<http://www.itksnap.org>). The rater was blinded regarding clinical and imaging data and followed a previously published protocol for ChP segmentation^15^. The ChP as well as the ventricle measures were adjusted for total intracranial volume using the proportions method^16^.

Structural MRI data quality control was performed as previously described by our working group^17^. Specifically, a visual inspection and utilization of the quality control software MRIQC^18^ were conducted. Manual assessment included rating overall image quality and evaluating specific sequence-related metrics such as signal-to-noise ratio (SNR), contrast-to-noise ratio (CNR), coefficient of joint variation (CJV), foreground-background energy ratio (FBER), median intensity non-uniformity (INU). Images were flagged if they received poor manual quality ratings and/or showed at least one abnormal quality metric. After image processing, we visually inspected the FreeSurfer segmentations. No participants were excluded based on criteria related to insufficient image quality.

The structural isotropic 3D T1-weighted MR images were processed with FreeSurfer v7.2, employing *recon-all*^17^. It included motion correction and averaging^19^, non-brain tissue removal^20^, automated Talairach transformation, segmentation of subcortical white and grey matter volumes^14,21^, intensity normalization^22^, tessellation of the grey matter-white matter boundary, automated topology correction^23,24^, and surface deformation^25,26,27^. More information regarding the FreeSurfer pipeline is available at <http://surfer.nmr.mgh.harvard.edu/>.

**Figure Legends**

**Figure S1.** **Loadings Plot for Principal Component Analysis of BCB permeability markers.**

Loadings plot illustrating the projection of variables on the first two principal components from a Principal Component Analysis (PCA). The variables included in the analysis are total CSF protein, CSF/serum albumin ratio, and CSF/serum IgG ratio. The first principal component (PC1) is plotted on the x-axis, and the second principal component (PC2) is plotted on the y-axis. The size of the bubbles represents the magnitude of the loadings. N = 57. Abbreviations: BCB, blood-cerebrospinal fluid barrier; CSF, cerebrospinal fluid; PC1, first principal component; PC2, second principal component.

**Figure S2.** **Relationship between blood-cerebrospinal fluid barrier integrity and measures of psychopathology as well as cognition.**

Regression plots illustrating associations between blood-cerebrospinal fluid barrier composite score, **(A)** PANSS positive, **(B)** PANSS negative, **(C)** PANSS general scores, **(D)** TMT-A, **(E)** TMT-B scores. Multiple univariate linear regression models were employed, controlling for age, sex and in (D) and (E) years of education. N = 47 in (A) – (C) and N = 34 in (D) and (E). Abbreviations: N, number of participants; BCB, blood-cerebrospinal fluid barrier; PANSS, Positive And Negative Syndrome Scale; TMT, Trail Making Test.

**Figure S3. Relationship between illness characteristics and blood-cerebrospinal fluid barrier integrity.**

Regression plots illustrating associations between **(A)** duration of illness and **(C)** duration of treatment and blood-cerebrospinal fluid barrier composite score. **(B)** Comparison of mean blood-cerebrospinal fluid barrier composite score between individuals with first-episode psychosis (red) and multiple-episode psychosis (turquoise) illustrated with box and violin plots. Multiple univariate linear regression models were employed, controlling for age and sex. N = 56 in (A), (C), and N = 57 in (B). Abbreviations: N, number of participants; BCB, blood-cerebrospinal fluid barrier; AP, antipsychotic; FEP, first-episode psychosis; MEP, multiple-episode psychosis.

**Figure S4. Relationship between blood-cerebrospinal fluid barrier integrity and cardiovascular factors.**

Regression plots illustrating associations between blood-cerebrospinal fluid barrier composite score, **(A)** glycated haemoglobin (HbA1c) and **(B)** systolic blood pressure. Two linear regression models were employed, controlling for age, sex, BMI and smoking status. N = 52 in (A) and N = 55 in (B). Abbreviations: N, number of participants; BCB, blood-cerebrospinal fluid barrier; HbA1c, glycated haemoglobin; systolic BP, systolic blood pressure.

**Figure S5. Relationship between blood-cerebrospinal fluid barrier integrity and inflammatory markers.**

Regression plots illustrating associations between blood-cerebrospinal fluid barrier composite score, **(A)** absolute lymphocyte count and **(B)** C-reactive protein. Two linear regression models were employed, controlling for age, sex, BMI and smoking status. N = 54. Abbreviations: N, number of participants; BCB, blood-cerebrospinal fluid barrier; C-reactive protein.

**Figure S6. Relationship between blood-cerebrospinal fluid barrier integrity and 3^rd^ or 4^th^ ventricle.**

Regression plots illustrating associations between blood-cerebrospinal fluid barrier composite score, **(A)** 3^rd^ ventricle volume and **(B)** 4^th^ ventricle volume. Multiple linear regression models were employed, controlling for age and sex. N = 28. Abbreviations: N, number of participants; BCB, blood-cerebrospinal fluid barrier.

**Tables**

**Table St1 CSF parameter characteristics.**

| **CSF parameters** | **SSD**  *Mean* ± *SD*  *n (%)* | | | *N* |
| --- | --- | --- | --- | --- |
| Protein level (mg/dl) | 42.42 ± 18.55 | | | 57 |
| Protein level elevated (yes:no) | 17:40 (29.8%) | | | 57 |
| CSF/serum albumin ratio | 6.53 ± 3.36 | | | 57 |
| CSF/serum albumin ratio elevated* (yes:no) | 24:33 (42.1%) | | | 57 |
| CSF/serum IgG ratio | 3.18 ± 1.66 | | | 57 |
| White Blood Cell Count (cells/µl) | 0.88 ± 1.09 | | | 57 |
| Pleocytosis (> 5/µl)) (yes:no) | 0:57 | | | 57 |
| OCBs (yes:no) | 5:52 (8.8%) | | | 57 |
| OCBs intrathecal synthesis (yes:no) | 0:57 | | | 57 |
| Neuronal autoantibodies** (yes:no) |  | 0:55 | 55 | |

CSF, cerebrospinal fluid; *N*, number of participants; OCBs, oligoclonal IgG bands; *SD*, standard deviation

*CSF/serum albumin ratio (Qalb) cut-off is adjusted for age according to the formula Qalb = (4 + age/15) × 10^–3^

**neuronal antibodies include NMDA antibodies, Glutamate (AMPA 1/2) receptor antibodies, DPPX antibodies, CASPR2 antibodies, LGI1 antibodies, GABA B (1/2) antibodies

**Table St2 Blood parameter characteristics.**

| **Blood Parameters** | **Patients with SSD**  *Mean* ± *SD*  *n (%)* | *N* |
| --- | --- | --- |
| Total cholesterol (mg/dl) | 178.20 ± 32.64 | 53 |
| Total cholesterol elevated (yes:no) | 14:41 (25.5%) | 53 |
| HDL cholesterol (mg/dl) | 53.24 ± 16.48 | 53 |
| HDL cholesterol reduced (yes:no) | 27:28 (49.1%) | 53 |
| LDL cholesterol (mg/dl) | 109.10 ± 37.74 | 53 |
| LDL cholesterol elevated (yes:no) | 19:36 (34.5%) | 53 |
| Triglyceride (mg/dl) | 118.50 ± 82.99 | 53 |
| Triglyceride elevated (yes:no) | 14:41 (25.5%) | 53 |
| HbA1c (%) | 5.34 ± 0.51 | 52 |
| HbA1c elevated (yes:no) | 3:51 (5.6%) | 52 |
| Serum-CRP (mg/dl) | 0.18 ± 0.26 | 54 |
| Serum-CRP elevated (yes:no) | 5:51 (8.9%) | 54 |
| Neutrophiles (thou./µl) | 4.27 ± 1.82 | 54 |
| Neutrophiles elevated (yes:no) | 3:53 (5.4%) | 54 |
| Monocytes (thou./µl) | 0.53 ± 0.17 | 54 |
| Monocytes elevated (yes:no) | 3:53 (5.4%) | 54 |
| Lymphocytes (thou./µl) | 1.86 ± 0.57 | 54 |
| Lymphocytes elevated (yes:no) | 0:56 (0%) | 54 |
| NLR | 2.43 ± 1.03 | 54 |
| MLR | 0.30 ± 0.10 | 54 |

MLR, Monocyte-Lymphocyte-Ratio; *N*, number of participants; NLR, Neutrophile-Lymphocyte-Ratio; *SD*, standard deviation

| **List of Abbreviations** | |
| --- | --- |
| Abbreviation | Explanation |
| BC | blood count |
| BCB | blood-cerebrospinal fluid-barrier |
| BMI | body-mass-index |
| CI | confidence intervall |
| CRP | C-reactive protein |
| CSF | cerebrospinal fluid |
| DUI | duration of illness |
| FEP | first episode psychosis |
| GAF | Global Assessment of Functioning |
| HDL | high-density lipoprotein |
| LDL | low-density lipoprotein |
| LL | lower limit |
| MLR | monocyte-to-lymphocyte ratio |
| MoCA | Montreal Cognitive Assessment |
| N | number of participants |
| NLR | neutrophile-to-lymphocyte ratio |
| p | p-value |
| PANSS | Positive And Negative Syndrome Scale |
| q | false dicovery rate adjustet p-value |
| SSD | schizophrenia-spectrum disorder |
| TMT | Trail-Making Test |
| UL | upper limit |

| **Table S1 Complete parameter estimates – association analyses of general disease characteristic measures and blood-CSF barrier (BCB) composite score in SSD** | | | | | | |
| --- | --- | --- | --- | --- | --- | --- |
| **Table S1.1 Association between duration of illness (DUI) and BCB composite score** | | | |  |  |  |
| Response | Predictor | Estimate | 95% CI [LL, UL] | N | *p* | *q* |
| BCB composite score | (Intercept) | -1.404 | [-2.586, -0.022] | 56 | 0.052 | 0.078 |
| BCB composite score | DUI (months) | -0.003 | [-0.007, 0.001] | 56 | 0.25 | 0.519 |
| BCB composite score | age | 0.05 | [0.014 , 0.085] | 56 | 0.022* | 0.033* |
| BCB composite score | sex | -0.396 | [-1.270, 0.477] | 56 | 0.451 | 0.451 |
| **Table S1.2 Association between duration of antipsychotic treatment and BCB composite score** | | | | |  |  |
| Response | Predictor | Estimate | 95% CI [LL, UL] | N | *p* | *q* |
| BCB composite score | (Intercept) | -1.504 | [-2.694, -0.315] | 56 | 0.039* | 0.078 |
| BCB composite score | Duration AP treatment (months) | -0.002 | [-0.007, 0.002] | 56 | 0.346 | 0.519 |
| BCB composite score | age | 0.051 | [0.015, 0.086] | 56 | 0.021* | 0.033* |
| BCB composite score | sex | -0.442 | [-1.332, 0.448] | 56 | 0.409 | 0.451 |
| **Table S1.3 Association between FEP status and BCB composite score** | | | |  |  |  |
| Response | Predictor | Estimate | 95% CI [LL, UL] | N | *p* | *q* |
| BCB composite score | (Intercept) | -1.177 | [-2.422, 0.069] | 57 | 0.12 | 0.12 |
| BCB composite score | FEP status | -0.287 | [-1.055, 0.482] | 57 | 0.535 | 0.535 |
| BCB composite score | age | 0.042 | [0.010, 0.075] | 57 | 0.034* | 0.034* |
| BCB composite score | sex | -0.378 | [-1.254, 0.498] | 57 | 0.473 | 0.102 |

| **Table S2 Complete parameter estimates – association analyses of disease severity measures and blood-CSF barrier (BCB) composite score in SSD** | | | | | | |
| --- | --- | --- | --- | --- | --- | --- |
| **Table S2.1 Association between BCB composite score and GAF score** | | | |  |  |  |
| Response | Predictor | Estimate | 95% CI [LL, UL] | N | *p* | *q* |
| GAF | (Intercept) | 42.919 | [33.155, 52.684] | 47 | <0.001*** | <0.001*** |
| GAF | BCB composite score | 0.604 | [-1.026, 2.234] | 47 | 0.537 | 0.806 |
| GAF | age | 0.166 | [-0.124, 0.455] | 47 | 0.341 | 0.409 |
| GAF | sex | -5.927 | [-12.414, 0.560] | 47 | 0.132 | 0.163 |
| **Table S2.2 Association between BCB composite score and treatment resistance** | | | | |  |  |
| Response | Predictor | Estimate | 95% CI [LL, UL] | N | *p* | *q* |
| Clozapine lifetime | (Intercept) | -9.63 | [-17.454, -1.799] | 55 | 0.043* | 0.043* |
| Clozapine lifetime | BCB composite score | 0.668 | [0.158, 1.177] | 55 | 0.031* | 0.186 |
| Clozapine lifetime | age | 0.028 | [-0.044, 0.101] | 55 | 0.518 | 0.583 |
| Clozapine lifetime | sex | -1.817 | [-4.683, 1.050] | 55 | 0.297 | 0.297 |
| Clozapine lifetime | smoking status | 0.065 | [-1.677, 1.806] | 55 | 0.951 | 0.951 |
| Clozapine lifetime | BMI | 0.233 | [-0.018, 0.483] | 55 | 0.126 | 0.126 |
| **Table S2.3 Association between BCB composite score and PANSS total score** | | | | |  |  |
| Response | Predictor | Estimate | 95% CI [LL, UL] | N | *p* | *q* |
| PANSS total score | (Intercept) | 68.26 | [57.592, 78.928] | 47 | <0.001*** | <0.001*** |
| PANSS total score | BCB composite score | -0.153 | [-2.000, 1.694] | 47 | 0.89 | 0.89 |
| PANSS total score | age | -0.237 | [-0.561, 0.086] | 47 | 0.225 | 0.396 |
| PANSS total score | sex | 8.698 | [1.306, 16.089] | 47 | 0.054 | 0.163 |
| **Table S2.4 Association between BCB composite score and PANSS positive score** | | | | |  |  |
| Response | Predictor | Estimate | 95% CI [LL, UL] | N | *p* | *q* |
| PANSS positive score | (Intercept) | 13.627 | [10.231, 17.023] | 47 | <0.001*** | <0.001*** |
| PANSS positive score | BCB composite score | 0.255 | [-0.333, 0.843] | 47 | 0.47 | 0.806 |
| PANSS positive score | age | 0.034 | [-0.069, 0.137] | 47 | 0.583 | 0.583 |
| PANSS positive score | sex | 2.129 | [-0.224, 4.482] | 47 | 0.136 | 0.163 |
| **Table S2.5 Association between BCB composite score and PANSS negative score** | | | | |  |  |
| Response | Predictor | Estimate | 95% CI [LL, UL] | N | *p* | *q* |
| PANSS negative score | (Intercept) | 20.086 | [15.440, 24.732] | 47 | <0.001*** | <0.001*** |
| PANSS negative score | BCB composite score | 0.157 | [-0.647, 0.961] | 47 | 0.744 | 0.89 |
| PANSS negative score | age | -0.165 | [-0.306, -0.024] | 47 | 0.055 | 0.33 |
| PANSS negative score | sex | 3.364 | [0.145, 6.583] | 47 | 0.086 | 0.163 |
| **Table S2.6 Association between BCB composite score and PANSS general score** | | | | |  |  |
| Response | Predictor | Estimate | 95% CI [LL, UL] | N | *p* | *q* |
| PANSS general score | (Intercept) | 34.506 | [29.159, 39.854] | 47 | <0.001*** | <0.001*** |
| PANSS general score | BCB composite score | -0.53 | [-1.456, 0.396] | 47 | 0.341 | 0.806 |
| PANSS general score | age | -0.109 | [-0.271, 0.053] | 47 | 0.264 | 0.396 |
| PANSS general score | sex | 3.383 | [-0.323, 7.088] | 47 | 0.132 | 0.163 |

| **Table S3 Complete parameter estimates – association analyses of cognitive measures and blood-CSF barrier (BCB) composite score in SSD** | | | | | | |
| --- | --- | --- | --- | --- | --- | --- |
| **Table S3.1 Association between BCB composite score and TMT A time** | | | |  |  |  |
| Response | Predictor | Estimate | 95% CI [LL, UL] | N | *p* | *q* |
| TMT A time | (Intercept) | 42.571 | [23.891, 61.250] | 34 | 0.001*** | 0.002** |
| TMT A time | BCB composite score | -0.527 | [-2.620, 1.566] | 34 | 0.672 | 0.672 |
| TMT A time | age | 0.552 | [0.131, 0.973] | 34 | 0.034* | 0.102 |
| TMT A time | sex | -3.983 | [-13.937, 5.972] | 34 | 0.502 | 0.864 |
| TMT A time | education (years) | -1.786 | [-2.882, -0.690] | 34 | 0.01** | 0.015* |
| **Table S3.2 Association between BCB composite score and TMT B time** | | | |  |  |  |
| Response | Predictor | Estimate | 95% CI [LL, UL] | N | *p* | *q* |
| TMT B time | (Intercept) | 143.756 | [53.514, 233.999] | 34 | 0.011* | 0.011* |
| TMT B time | BCB composite score | -5.215 | [-15.328,  4.898] | 34 | 0.388 | 0.672 |
| TMT B time | age | 0.973 | [-1.060, 3.006] | 34 | 0.423 | 0.423 |
| TMT B time | sex | -4.759 | [-52.851, 43.333] | 34 | 0.868 | 0.868 |
| TMT B time | education (years) | -5.37 | [-10.667, -0.074] | 34 | 0.096 | 0.096 |
| **Table S3.3 Association between BCB composite score and MoCA score** | | | |  |  |  |
| Response | Predictor | Estimate | 95% CI [LL, UL] | N | *p* | *q* |
| MoCA score | (Intercept) | 18.786 | [14.282, 23.290] | 35 | <0.001*** | <0.001*** |
| MoCA score | BCB composite score | 0.185 | [-0.319, 0.689] | 35 | 0.538 | 0.672 |
| MoCA score | age | -0.078 | [-0.179, 0.023] | 35 | 0.2 | 0.3 |
| MoCA score | sex | -0.799 | [-3.196, 1.599] | 35 | 0.576 | 0.864 |
| MoCA score | education (years) | 0.623 | [0.361, 0.886] | 35 | <0.001*** | 0.001*** |

| **Table S4 Complete parameter estimates – association analyses of cardiometabolic factors and blood-CSF barrier (BCB) composite score in SSD** | | | | | | |
| --- | --- | --- | --- | --- | --- | --- |
| **Table S4.1 Association between BCB composite score and total cholesterol** | | | | |  |  |
| Response | Predictor | Estimate | 95% CI [LL, UL] | N | *p* | *q* |
| BCB composite score | (Intercept) | -2.03 | [-4.511, 0.452] | 53 | 0.177 | 0.212 |
| BCB composite score | total cholesterol | 0.026 | [0.014, 0.038] | 53 | 0.001** | 0.001** |
| BCB composite score | age | 0.031 | [-0.002, 0.064] | 53 | 0.121 | 0.145 |
| BCB composite score | sex | -0.381 | [-1.175, 0.412] | 53 | 0.424 | 0.761 |
| BCB composite score | smoker status | 0.679 | [-0.010, 1.368] | 53 | 0.105 | 0.152 |
| BCB composite score | BMI | -0.147 | [-0.237, -0.056] | 53 | 0.009** | 0.022* |
| **Table S4.2 Association between BCB composite score and LDL cholesterol** | | | | |  |  |
| Response | Predictor | Estimate | 95% CI [LL, UL] | N | *p* | *q* |
| BCB composite score | (Intercept) | -0.006 | [-2.293, 2.282] | 53 | 0.997 | 0.997 |
| BCB composite score | LDL cholesterol | 0.023 | [0.013, 0.033] | 53 | <0.001*** | 0.001** |
| BCB composite score | age | 0.027 | [-0.006, 0.060] | 53 | 0.182 | 0.182 |
| BCB composite score | sex | -0.236 | [-1.026, 0.553] | 53 | 0.618 | 0.761 |
| BCB composite score | smoker status | 0.807 | [0.124, 1.489] | 53 | 0.053 | 0.152 |
| BCB composite score | BMI | -0.14 | [-0.229, -0.051] | 53 | 0.011* | 0.022* |
| **Table S4.3 Association between BCB composite score and HDL cholesterol** | | | | |  |  |
| Response | Predictor | Estimate | 95% CI [LL, UL] | N | *p* | *q* |
| BCB composite score | (Intercept) | 3.579 | [0.619, 6.538] | 53 | 0.048* | 0.212 |
| BCB composite score | HDL cholesterol | -0.045 | [-0.067, -0.022] | 53 | 0.002** | 0.003** |
| BCB composite score | age | 0.053 | [0.021,  0.085] | 53 | 0.008 | 0.024* |
| BCB composite score | sex | -0.02 | [-0.842, 0.802] | 53 | 0.968 | 0.968 |
| BCB composite score | smoker status | 0.613 | [-0.094, 1.321] | 53 | 0.152 | 0.152 |
| BCB composite score | BMI | -0.123 | [-0.212, -0.033] | 53 | 0.026* | 0.039* |
| **Table S4.4 Association between BCB composite score and triglycerides** | | | | |  |  |
| Response | Predictor | Estimate | 95% CI [LL, UL] | N | *p* | *q* |
| BCB composite score | (Intercept) | 1.803 | [-0.393, 4.000] | 53 | 0.175 | 0.212 |
| BCB composite score | Triglycerides | 0.013 | [0.008, 0.017] | 53 | <0.001*** | <0.001*** |
| BCB composite score | age | 0.058 | [0.029, 0.087] | 53 | 0.002** | 0.012* |
| BCB composite score | sex | -0.218 | [-0.947, 0.511] | 53 | 0.619 | 0.761 |
| BCB composite score | smoker status | 0.666 | [0.034, 1.297] | 53 | 0.084 | 0.152 |
| BCB composite score | BMI | -0.209 | [-0.299, -0.120] | 53 | <0.001*** | 0.002** |
| **Table S4.5 Association between BCB composite score and systolic blood pressure** | | | | |  |  |
| Response | Predictor | Estimate | 95% CI [LL, UL] | N | *p* | *q* |
| BCB composite score | (Intercept) | -4.027 | [-8.622, 0.567] | 55 | 0.148 | 0.212 |
| BCB composite score | systolic BP | 0.037 | [0.001, 0.073] | 55 | 0.088 | 0.105 |
| BCB composite score | age | 0.04 | [0.007, 0.074] | 55 | 0.05* | 0.077 |
| BCB composite score | sex | -0.249 | [-1.121, 0.623] | 55 | 0.634 | 0.761 |
| BCB composite score | smoker status | 0.656 | [-0.082, 1.395] | 55 | 0.143 | 0.152 |
| BCB composite score | BMI | -0.081 | [-0.174, 0.011] | 55 | 0.146 | 0.146 |
| **Table S4.6 Association between BCB composite score and HbA1c** | | | |  |  |  |
| Response | Predictor | Estimate | 95% CI [LL, UL] | N | *p* | *q* |
| BCB composite score | (Intercept) | -3.441 | [-7.607, 0.726] | 52 | 0.172 | 0.212 |
| BCB composite score | HbA1c | 0.819 | [-0.012, 1.650] | 52 | 0.105 | 0.105 |
| BCB composite score | age | 0.044 | [0.007, 0.080] | 52 | 0.051 | 0.077 |
| BCB composite score | sex | -0.381 | [-1.258, 0.496] | 52 | 0.469 | 0.761 |
| BCB composite score | smoker status | 0.818 | [0.052, 1.583] | 52 | 0.08 | 0.152 |
| BCB composite score | BMI | -0.1 | [-0.198, -0.002] | 52 | 0.095 | 0.114 |

| **Table S5 Complete parameter estimates – association analyses of peripheral inflammatory markers and blood-CSF barrier (BCB) composite score in SSD** | | | | | | |
| --- | --- | --- | --- | --- | --- | --- |
| **Table S5.1 Association between BCB composite score and absolute neutrophil count** | | | | |  |  |
| Response | Predictor | Estimate | 95% CI [LL, UL] | N | *p* | *q* |
| BCB composite score | (Intercept) | 0.238 | [-2.329, 2.805] | 54 | 0.87 | 1 |
| BCB composite score | BC neutrophil granulocytes | 0.136 | [-0.095, 0.367] | 54 | 0.328 | 0.949 |
| BCB composite score | age | 0.045 | [0.011, 0.079] | 54 | 0.033* | 0.039* |
| BCB composite score | sex | -0.529 | [-1.431, 0.373] | 54 | 0.331 | 0.443 |
| BCB composite score | smoker status | 0.575 | [-0.234, 1.383] | 54 | 0.239 | 0.271 |
| BCB composite score | BMI | -0.093 | [-0.191, 0.006] | 54 | 0.122 | 0.21 |
| **Table S5.2 Association between BCB composite score and absolute monocyte count** | | | | |  |  |
| Response | Predictor | Estimate | 95% CI [LL, UL] | N | *p* | *q* |
| BCB composite score | (Intercept) | 0 | [-2.651, 2.650] | 54 | 1 | 1 |
| BCB composite score | BC monocytes | 1.25 | [-1.287, 3.787] | 54 | 0.413 | 0.949 |
| BCB composite score | age | 0.045 | [0.011, 0.079] | 54 | 0.031* | 0.039* |
| BCB composite score | sex | -0.492 | [-1.389, 0.406] | 54 | 0.363 | 0.443 |
| BCB composite score | smoker status | 0.559 | [-0.282, 1.400] | 54 | 0.271 | 0.271 |
| BCB composite score | BMI | -0.088 | [-0.186, 0.010] | 54 | 0.138 | 0.21 |
| **Table S5.3 Association between BCB composite score and absolute lymphocyte count** | | | | |  |  |
| Response | Predictor | Estimate | 95% CI [LL, UL] | N | *p* | *q* |
| BCB composite score | (Intercept) | 0.303 | [-2.315, 2.920] | 54 | 0.847 | 1 |
| BCB composite score | BC lymphocytes | 0.03 | [-0.755, 0.816] | 54 | 0.949 | 0.949 |
| BCB composite score | age | 0.046 | [0.011, 0.081] | 54 | 0.031* | 0.039* |
| BCB composite score | sex | -0.426 | [-1.336, 0.485] | 54 | 0.437 | 0.443 |
| BCB composite score | smoker status | 0.721 | [-0.085, 1.528] | 54 | 0.14 | 0.21 |
| BCB composite score | BMI | -0.081 | [-0.189, 0.026] | 54 | 0.209 | 0.21 |
| **Table S5.4 Association between BCB composite score and NLR** | | |  |  |  |  |
| Response | Predictor | Estimate | 95% CI [LL, UL] | N | *p* | *q* |
| BCB composite score | (Intercept) | 0.002 | [-2.755, 2.759] | 54 | 0.999 | 1 |
| BCB composite score | NLR | 0.133 | [-0.276, 0.541] | 54 | 0.588 | 0.949 |
| BCB composite score | age | 0.044 | [0.009, 0.079] | 54 | 0.039* | 0.039* |
| BCB composite score | sex | -0.429 | [-1.318, 0.460] | 54 | 0.423 | 0.443 |
| BCB composite score | smoker status | 0.705 | [-0.068, 1.478] | 54 | 0.132 | 0.21 |
| BCB composite score | BMI | -0.076 | [-0.174, 0.021] | 54 | 0.195 | 0.21 |
| **Table S5.5 Association between BCB composite score and MLR** | | |  |  |  |  |
| Response | Predictor | Estimate | 95% CI [LL, UL] | N | *p* | *q* |
| BCB composite score | (Intercept) | 0.035 | [-2.996, 3.066] | 54 | 0.984 | 1 |
| BCB composite score | MLR | 0.745 | [-3.425, 4.914] | 54 | 0.766 | 0.949 |
| BCB composite score | age | 0.045 | [0.010, 0.080] | 54 | 0.035* | 0.039* |
| BCB composite score | sex | -0.411 | [-1.303, 0.480] | 54 | 0.443 | 0.443 |
| BCB composite score | smoker status | 0.705 | [-0.079, 1.489] | 54 | 0.138 | 0.21 |
| BCB composite score | BMI | -0.076 | [-0.175, 0.024] | 54 | 0.209 | 0.21 |
| **Table S5.6 Association between BCB composite score and CRP** | | |  |  |  |  |
| Response | Predictor | Estimate | 95% CI [LL, UL] | N | *p* | *q* |
| BCB composite score | (Intercept) | 0.366 | [-2.430, 3.163] | 54 | 0.827 | 1 |
| BCB composite score | serum CRP | 0.081 | [-1.666, 1.828] | 54 | 0.939 | 0.949 |
| BCB composite score | age | 0.046 | [0.011, 0.080] | 54 | 0.03* | 0.039* |
| BCB composite score | sex | -0.43 | [-1.358, 0.497] | 54 | 0.44 | 0.443 |
| BCB composite score | smoker status | 0.737 | [-0.049, 1.524] | 54 | 0.122 | 0.21 |
| BCB composite score | BMI | -0.082 | [-0.190, 0.026] | 54 | 0.21 | 0.21 |

| **Table S6 Complete parameter estimates – association analyses of cerebroventricular measures and blood-CSF barrier (BCB) composite score in SSD** | | | | | | |
| --- | --- | --- | --- | --- | --- | --- |
| **Table S6.1 Association between BCB composite score and left choroid plexus volume** | | | | |  |  |
| Response | Predictor | Estimate | 95% CI [LL, UL] | N | *p* | *q* |
| Left choroid plexus | (Intercept) | 903.942 | [688.902, 1118.981] | 28 | <0.001 | <0.001 |
| Left choroid plexus | BCB composite score | 10.038 | [-23.318, 43.393] | 28 | 0.611 | 0.941 |
| Left choroid plexus | age | -5.64 | [-12.065, 0.785] | 28 | 0.146 | 0.468 |
| Left choroid plexus | sex | 7.911 | [-175.489, 191.310] | 28 | 0.942 | 0.942 |
| **Table S6.2 Association between BCB composite score and right choroid plexus volume** | | | | |  |  |
| Response | Predictor | Estimate | 95% CI [LL, UL] | N | *p* | *q* |
| Right choroid plexus | (Intercept) | 819.797 | [638.423, 1001.172] | 28 | <0.001 | <0.001 |
| Right choroid plexus | BCB composite score | 8.084 | [-20.050, 36.218] | 28 | 0.627 | 0.941 |
| Right choroid plexus | age | -4.085 | [-9.504, 1.334] | 28 | 0.209 | 0.468 |
| Right choroid plexus | sex | -82.558 | [-237.247, 72.130] | 28 | 0.37 | 0.942 |
| **Table S6.3 Association between BCB composite score and left lateral ventricle volume** | | | | |  |  |
| Response | Predictor | Estimate | 95% CI [LL, UL] | N | *p* | *q* |
| Left lateral ventricle | (Intercept) | 10419.46 | [6577.246, 14261.675] | 28 | <0.001 | <0.001 |
| Left lateral ventricle | BCB composite score | -47.78 | [-643.763, 548.202] | 28 | 0.892 | 0.984 |
| Left lateral ventricle | age | -56.766 | [-171.561, 58.029] | 28 | 0.406 | 0.491 |
| Left lateral ventricle | sex | -1174.105 | [-4450.994, 2102.783] | 28 | 0.546 | 0.942 |
| **Table S6.4 Association between BCB composite score and right lateral ventricle volume** | | | | |  |  |
| Response | Predictor | Estimate | 95% CI [LL, UL] | N | *p* | *q* |
| Right lateral ventricle | (Intercept) | 9038.154 | [6191.923, 11884.384] | 28 | <0.001 | <0.001 |
| Right lateral ventricle | BCB composite score | -183.642 | [-625.133, 257.849] | 28 | 0.484 | 0.941 |
| Right lateral ventricle | age | -40.696 | [-125.734, 44.341] | 28 | 0.421 | 0.491 |
| Right lateral ventricle | sex | -804.122 | [-3231.571, 1623.327] | 28 | 0.576 | 0.942 |
| **Table S6.5 Association between BCB composite score and third ventricle volume** | | | |  |  |  |
| Response | Predictor | Estimate | 95% CI [LL, UL] | N | *p* | *q* |
| 3rd ventricle | (Intercept) | 1230.933 | [796.736, 1665.131] | 28 | <0.001 | <0.001 |
| 3rd ventricle | BCB composite score | 36.555 | [-30.795, 103.905] | 28 | 0.362 | 0.941 |
| 3rd ventricle | age | -5.308 | [-18.280, 7.665] | 28 | 0.491 | 0.491 |
| 3rd ventricle | sex | -36.182 | [-406.493, 334.13] | 28 | 0.869 | 0.942 |
| **Table S6.6 Association between BCB composite score and fourth ventricle volume** | | | | |  |  |
| Response | Predictor | Estimate | 95% CI [LL, UL] | N | *p* | *q* |
| 4th ventricle | (Intercept) | 2516.088 | [1793.669, 3238.507] | 28 | <0.001 | <0.001 |
| 4th ventricle | BCB composite score | 1.348 | [-110.709, 113.406] | 28 | 0.984 | 0.984 |
| 4th ventricle | age | -15.406 | [-36.990, 6.178] | 28 | 0.234 | 0.468 |
| 4th ventricle | sex | -71.936 | [-688.061, 544.190] | 28 | 0.843 | 0.942 |
